# The effect of combining antibiotics on resistance: A systematic review and meta-analysis

**DOI:** 10.1101/2023.07.10.23292374

**Authors:** Berit Siedentop, Viacheslav N. Kachalov, Christopher Witzany, Matthias Egger, Roger D. Kouyos, Sebastian Bonhoeffer

## Abstract

When and under which conditions antibiotic combination therapy decelerates rather than accelerates resistance evolution is not well understood. We examined the effect of combining antibiotics on within-patient resistance development across various bacterial pathogens and antibiotics.

We searched CENTRAL, EMBASE and PubMed for (quasi)-randomised controlled trials (RCTs) published from database inception to November 24^th^, 2022. Trials comparing antibiotic treatments with different numbers of antibiotics were included. A patient was considered to have acquired resistance if, at the follow-up culture, a resistant bacterium (as defined by the study authors) was detected that had not been present in the baseline culture. We combined results using a random effects model and performed meta-regression and stratified analyses. The trials’ risk of bias was assessed with the Cochrane tool.

42 trials were eligible and 29, including 5054 patients, were qualified for statistical analysis. In most trials, resistance development was not the primary outcome and studies lacked power. The combined odds ratio (OR) for the acquisition of resistance comparing the group with the higher number of antibiotics with the comparison group was 1.23 (95% CI 0.68-2.25), with substantial between-study heterogeneity (*I^2^* =77%). We identified tentative evidence for potential beneficial or detrimental effects of antibiotic combination therapy for specific pathogens or medical conditions.

The evidence for combining a higher number of antibiotics compared to fewer from RCTs is scarce and overall, is compatible with both benefit or harm. Trials powered to detect differences in resistance development or well-designed observational studies are required to clarify the impact of combination therapy on resistance.

## Introduction

Antibiotics are one of the most significant advances in modern medicine, prescribed to treat various bacterial infections in both humans and animals and prevent infections, such as surgical site infections or opportunistic infections in immunocompromised individuals (1). However, this medical breakthrough is at risk due to the rising prevalence of antibiotic resistance and an inadequate pipeline of new antibiotics. This disturbing trend threatens to undermine the effectiveness of antibiotics and poses a severe challenge to public health worldwide (2, 3). Hence, we need a more prudent use of antibiotics, and where antibiotics are needed, we need treatment strategies that reduce the risk that resistance emerges or spreads. Different strategies for the optimal use of antibiotics have been investigated theoretically and empirically (4–7). Antibiotic combination therapy, i.e., the simultaneous administration of several antibiotics, is frequently discussed as a promising strategy for avoiding resistance evolution (6–10). Importantly, it is the standard of care for some bacterial pathogens, such as *H. pylori*, *Mycobacterium tuberculosis* (Mtb), or *Mycobacterium leprae* (11–13). However, it is unclear whether the effect of combination therapy on resistance is consistent for different pathogens.

There are several motivations for the use of antibiotic combination therapy, including to broaden the antibiotic spectrum in empirical treatment and reducing antibiotic resistance development (14, 15). The simultaneous occurrence of resistance mutations to multiple drugs is less likely than resistance to single drugs. Combination therapy should, therefore, reduce the development of resistance (10). This expectation is supported by viral infections such as HIV, where multiple point mutations are required for resistance to combination antiviral therapy. However, it is less clear to what extent this reasoning extends to antibiotic therapy, where the same mechanism can facilitate bacterial survival against multiple antibiotics (16, 17), and where horizontal transfer of resistance may occur. Indeed, the benefit of combining antibiotics for reducing resistance is debated for bacterial infections (18). Using more antibiotics overall could lead to more resistance, as overall antibiotic consumption correlates with resistance (19).

Two meta-analyses of randomised controlled trials (RCTs) comparing beta-lactam monotherapy to beta-lactam and aminoglycoside combination therapy found no differences in resistance development (4, 5). However, the effect of combining antibiotics on within-patient resistance development across many bacterial pathogens and various antibiotic combinations has not been addressed. Within-patient antibiotic resistance development, even if rare, may contribute to the emergence and spread of resistance. We performed a systematic review and meta-analysis to (i) test the effect of antibiotic combination therapy on within-patient resistance development and (ii) evaluate which factors affect the performance of combination therapy, as e.g. pathogen identity, treatment design and resistance assessment.

## Results

The search identified 3082 articles, which decreased to 1837 after deduplication. A total of 488 studies were eligible for full-text review, of which 41 studies qualified for inclusion. The screening of the citations of the 41 studies identified one additional eligible study (SI section 11.4), for a total of 42 studies, 40 RCTs and two quasi-RCTs, where the allocation method used is not truly random (figure 1, table 1) (20–61). Twenty-nine studies could be included in the meta-analysis; 13 were excluded due to zero events in both treatment arms.

**Figure 1.**
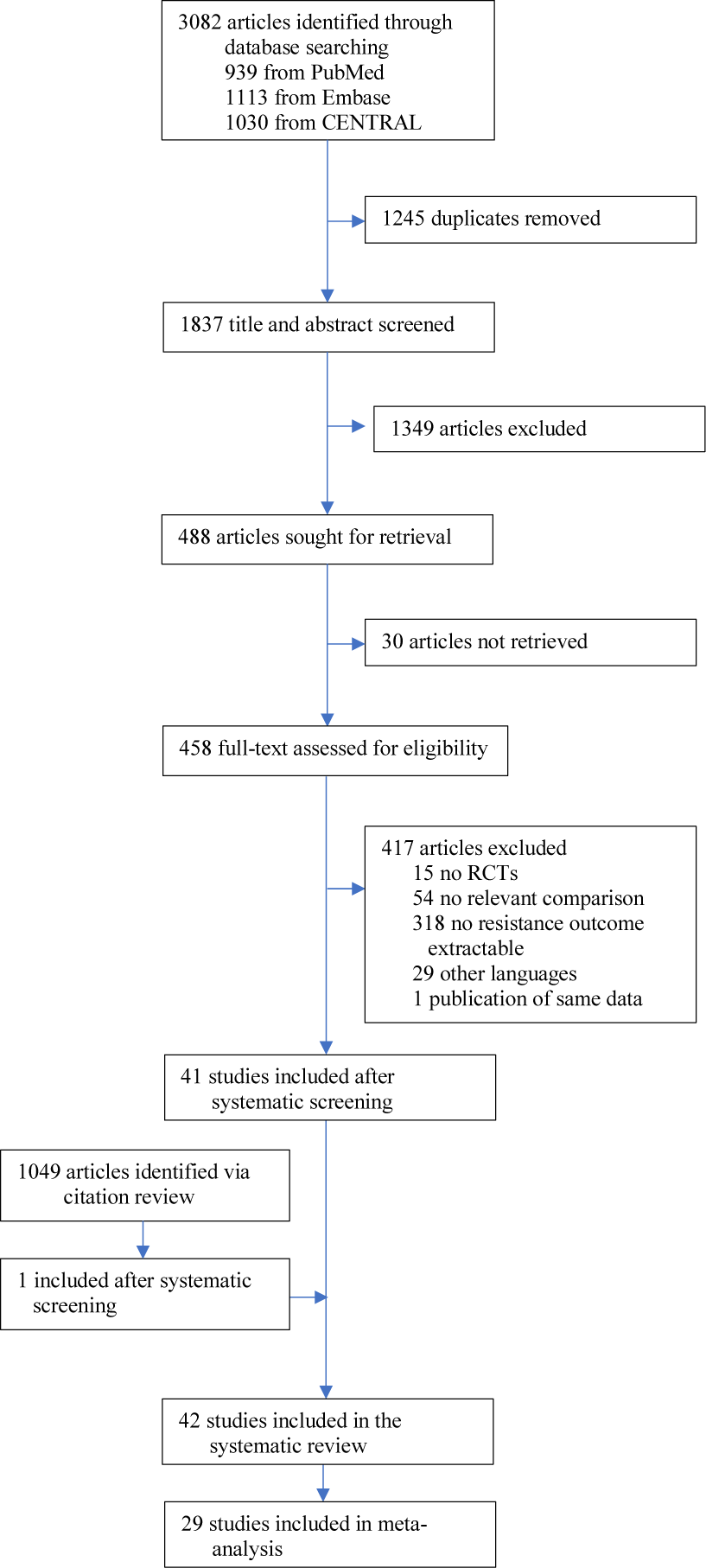
Study selection.

**Table 1.**
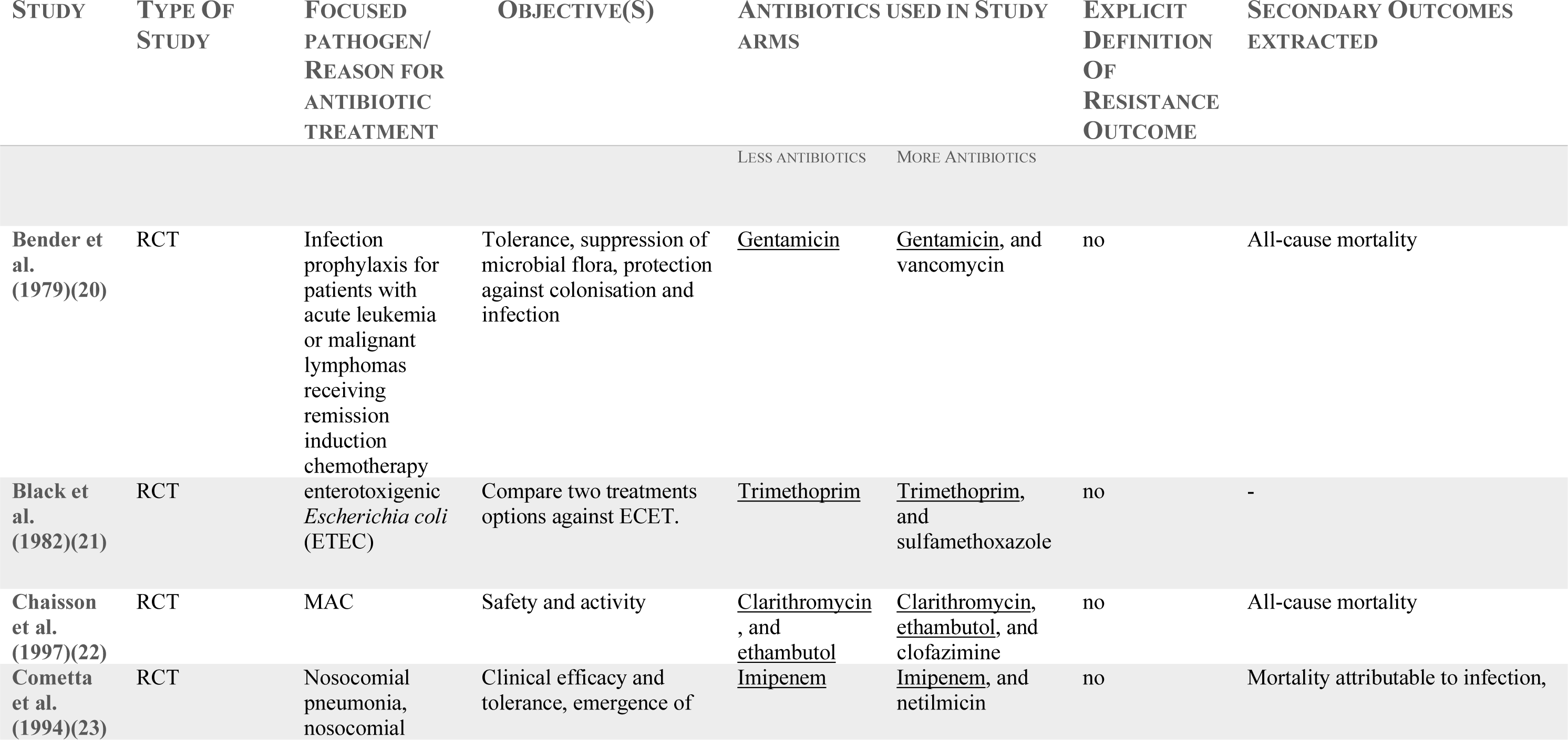
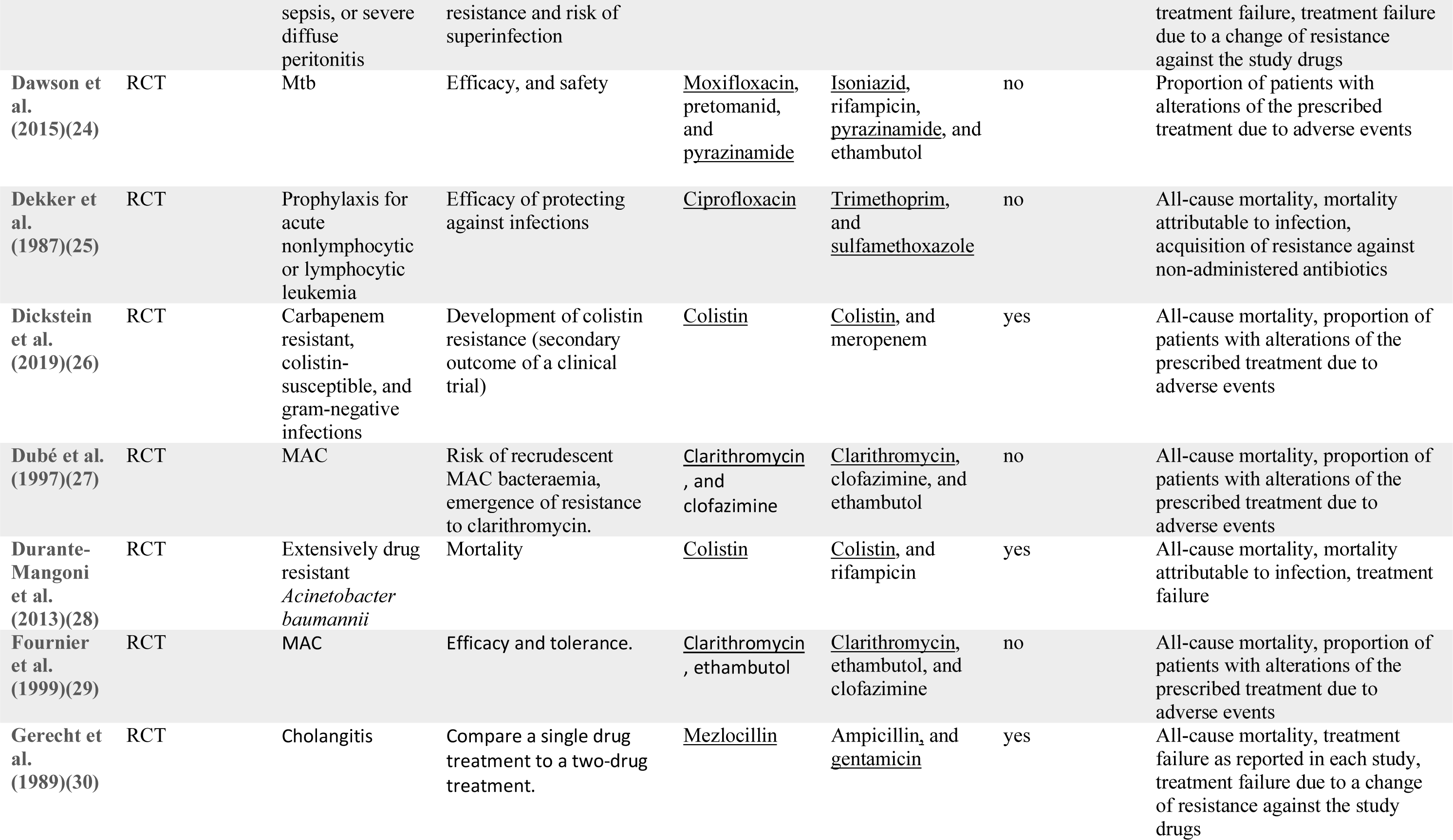
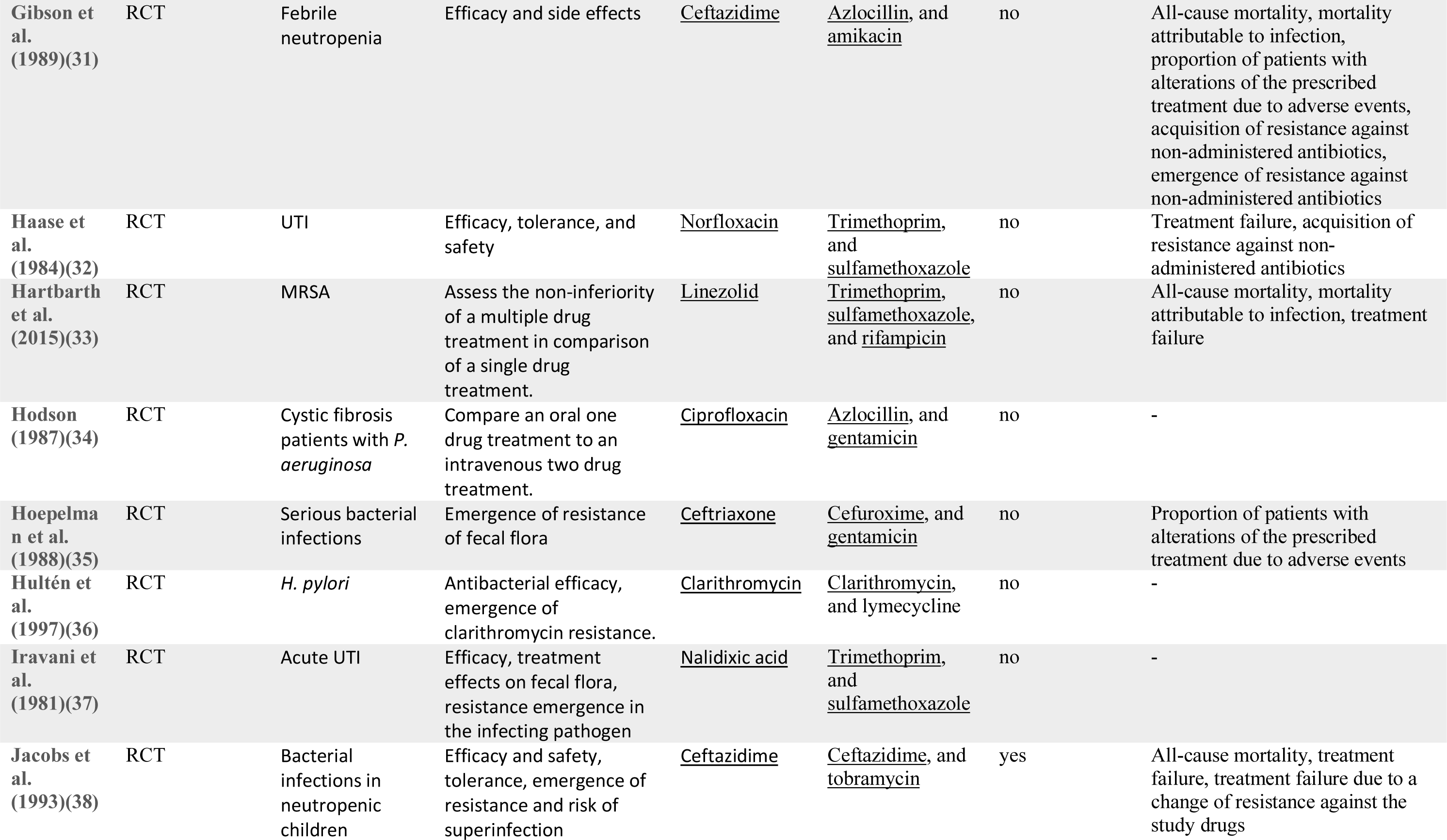
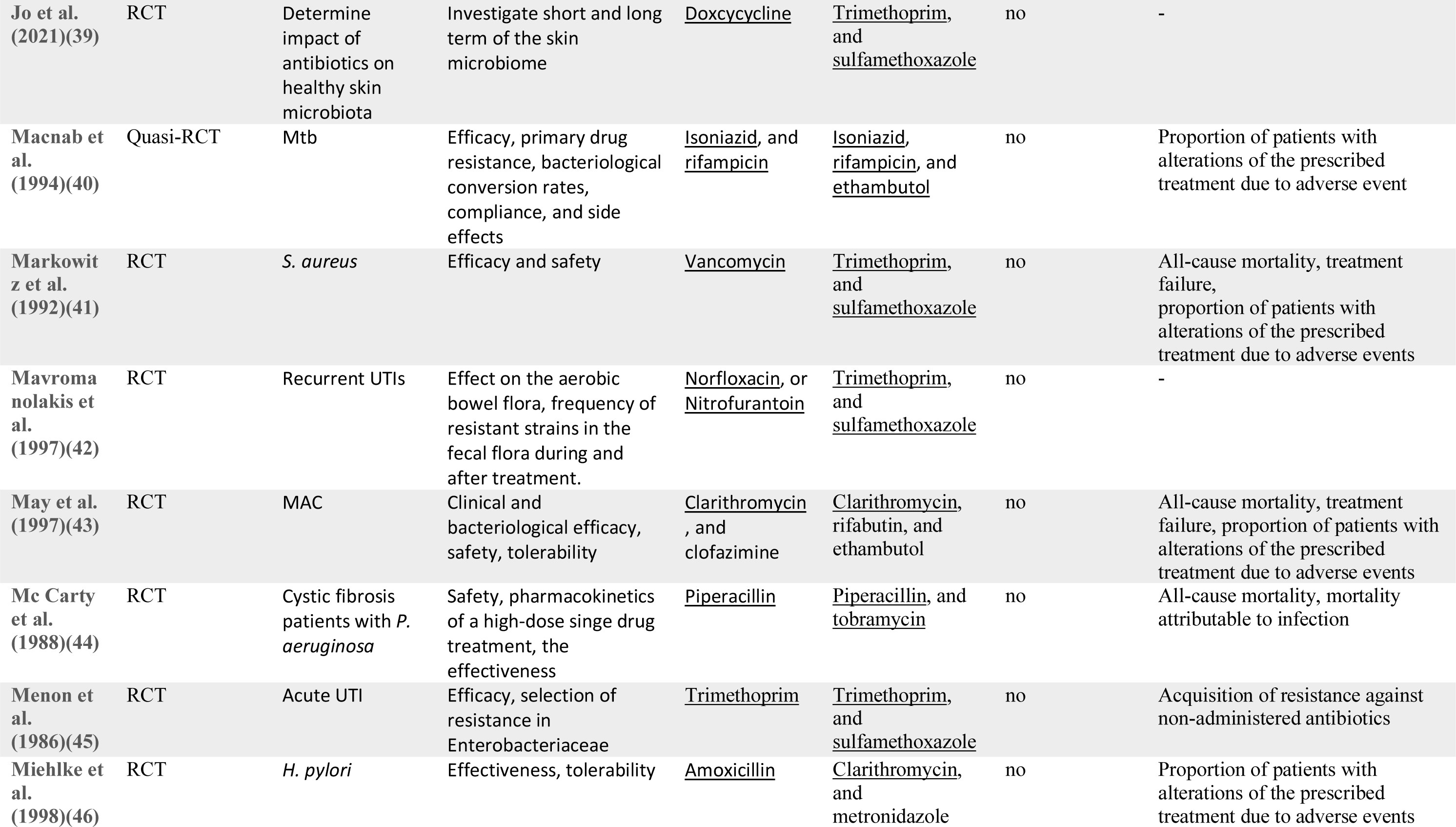
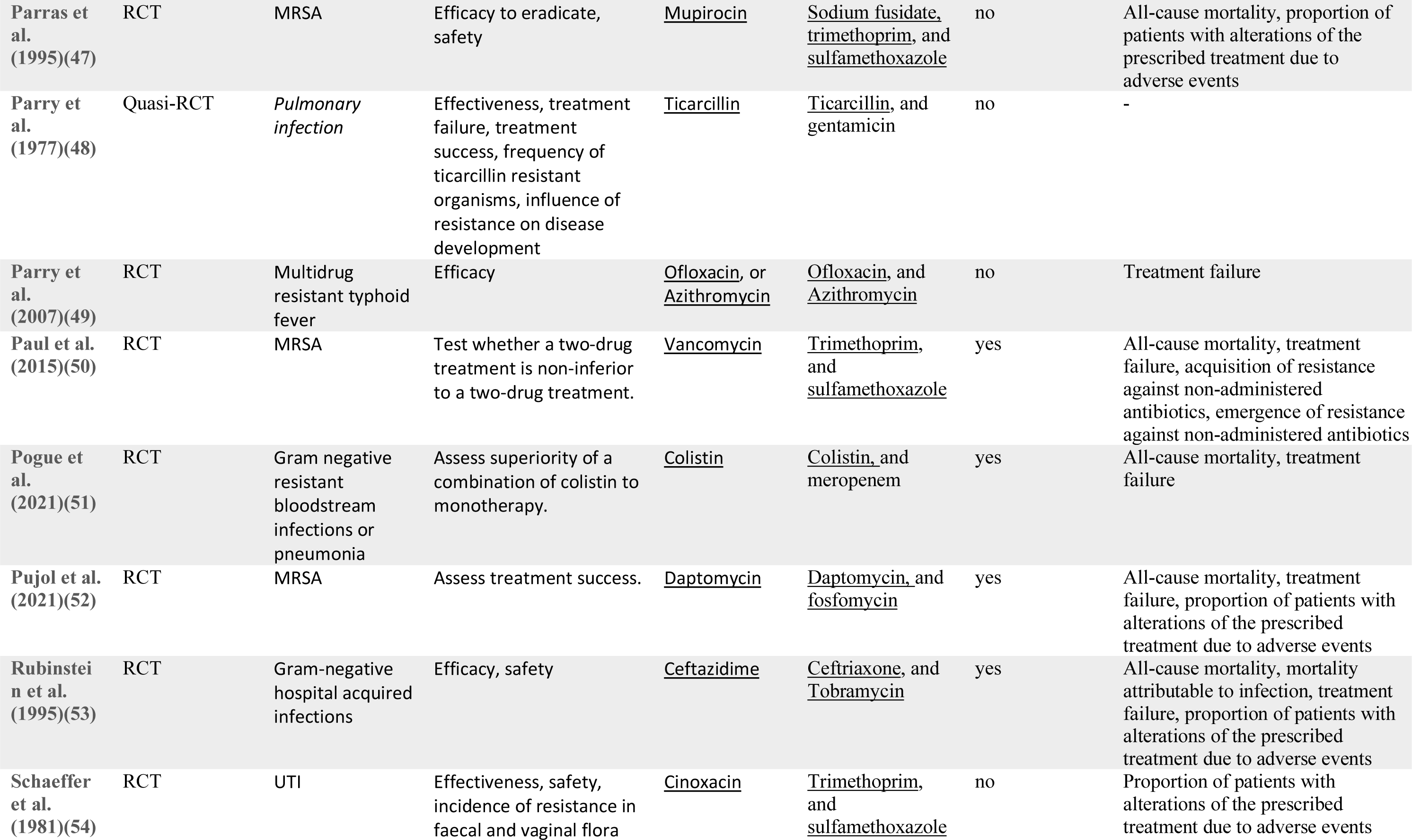
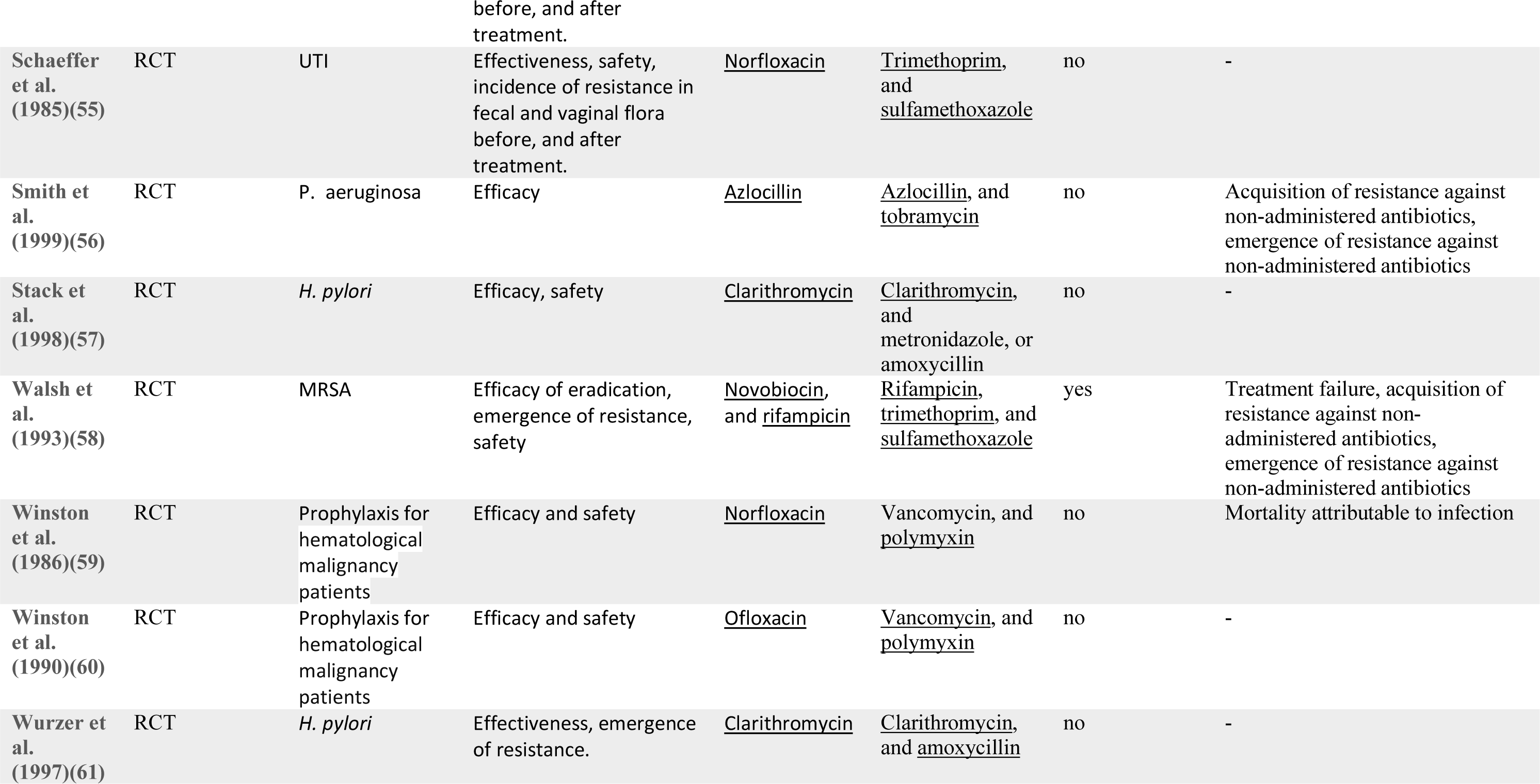
Overview of the 42 RCTs or quasi-RCTs included in the systematic review and meta-analysis. The underlined antibiotics indicate that resistance measurements were made for this antibiotic, reported and extractable from the studies. Justification for resistance outcome extraction is given in SI table S1.

The included studies were published between 1977 and 2021, with a median publication year of 1995 and few recent studies (figure 2 A). The development of antibiotic resistance was typically not the main outcome: only nine studies (21%) explicitly defined a resistance outcome (table 1, SI table S1). Consequently, most studies did not have the statistical power to detect differences in within-patient resistance development even if we assume that the effect on resistance development is large between treatment arms (figure 2 B, SI section 8). Twenty-two (52%) focused on a specific pathogen species (resistant *Acinetobacter baumannii*, *Escherichia coli*, *H. pylori*, Mtb, methicillin-resistant *Staphylococcus aureus* (MRSA), *Pseudomonas aeruginosa*, *Staphylococcus aureus*) or pathogen group (MAC, *Salmonella enterica* subsp. *enterica* serotype Thyphi, or *Salmonella enterica* subsp. *enterica* serotype Parthypi A).

**Figure 2.**
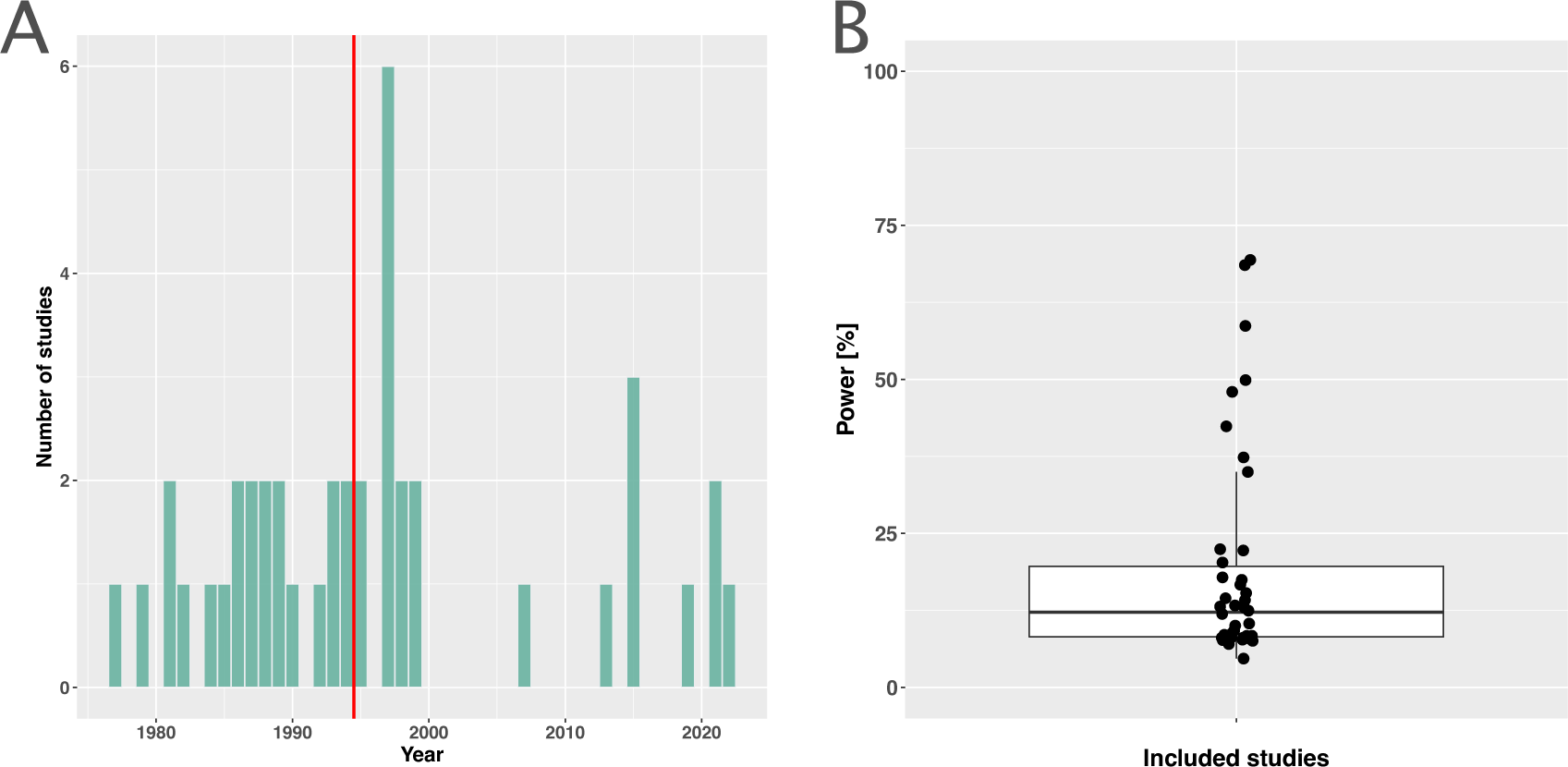
Measuring antibiotic resistance is not a current main objective of RCTs. A) Distribution of the publishing year of included studies, where n indicates the number of studies, and the red vertical line the median of the distribution. B) Calculated power of included studies to detect an odds ratio of 0.5. The power calculations were based on equal treatment arm sizes. For the calculations the treatment arm with the higher number of patients of the respective studies was used.

The five most frequent reasons for antibiotic administration were treatment or prophylaxis of urinary tract infections (UTIs) (6 studies, 14%), MRSA (5 studies, 12%), *H.* pylori, MAC, and prophylaxis for hematological malignancy patients with four studies (10%) respectively. Twenty-three of the included studies (55%) compared treatment arms with at least one administered antibiotic in common; the remaining studies compared treatment arms with no overlap in administered antibiotics (table 1). For the outcome acquisition of resistance, only two of all 42 studies had a low overall risk of bias according to the risk of bias assessment. Twelve (29%) were at high risk of bias, 28 (67%) at moderate risk of bias (SI section 3).

The overall pooled OR for acquisition of resistance comparing a lower number of antibiotics versus a higher one was 1.23 (95% CI 0.68 – 2.25), with substantial heterogeneity between studies (*I^2^* =77.4%). The latter OR was compatible with the OR for *de novo* emergence of resistance (pooled OR 0.74, 95% CI 0.34 – 1.59; *I^2^*=77%). The overall pooled estimates are based on studies that focus on various clinical conditions/pathogens and compare different antibiotics treatments. To explore the impact of these and other potential sources of heterogeneity on the resistance estimates we performed sub-group analyses and meta-regression. The results for the two resistance outcomes are qualitatively comparable in the sense that individual estimates may differ, but show overall similar absence of evidence to support either benefit, harm or equivalence of treating with a higher number of antibiotics. Therefore, our focus in the following is on the acquisition of resistance (details on emergence of resistance can be found in the SI sections 1-8).

Stratified analyses revealed that a higher number of antibiotics performed better than a lower number in case of *H. pylori*, (pooled OR 0.14, 95% CI 0.03 – 0.55; *I^2^* =41.7%, figure 3A), and *MAC* (pooled OR 0.18, 95% CI 0.06 – 0.52; *I^2^* =26.8%, figure 3A), but worse in case of *P. aeruginosa* (pooled OR 3.42, 95% CI 1.03 – 11.43; *I^2^*=1.54%, figure 3A). Furthermore, a lower number of antibiotics performed better than a higher number if the compared treatment arms had no antibiotics in common (pooled OR 4.73, 95% CI 2.14 – 10.42; *I^2^*=37%, SI table S3), which could be due to different potencies or resistance prevalences of antibiotics as discussed in SI (SI section 6.1.10). In contrast, when restricting the analysis to studies with at least one common antibiotic in the treatment arms we found no evidence of a difference, only a weak indication that a higher number of antibiotics performs better (pooled OR 0.55, 95% CI 0.28 – 1.07; *I^2^* =74%, figure 3B). When considering only resistance measurements of antibiotics common to both treatment arms instead of all resistance measurements, the arm with a higher number of antibiotics shows a benefit in comparison to the one with fewer (pooled OR 0.39, 95% CI 0.18 – 0.81; *I^2^*=75%, SI p 6). If the study measured the acquisition of resistance of both gram negative and positive bacteria, fewer antibiotics performed better (pooled OR 3.38, 95% CI 1.08 – 10.58; *I^2^*=38.35%, SI p 5). Other sub-group analyses did not show any harm or benefit of using a higher number of antibiotics. The results for all subgroup analyses are presented in the supplement (SI section 6). The multi-model inference for our meta-regression showed that the only significant factor influencing the outcome acquisition of resistance is whether at least one common antibiotic was used in the comparator arms (for details see SI section 7).

**Figure 3.**
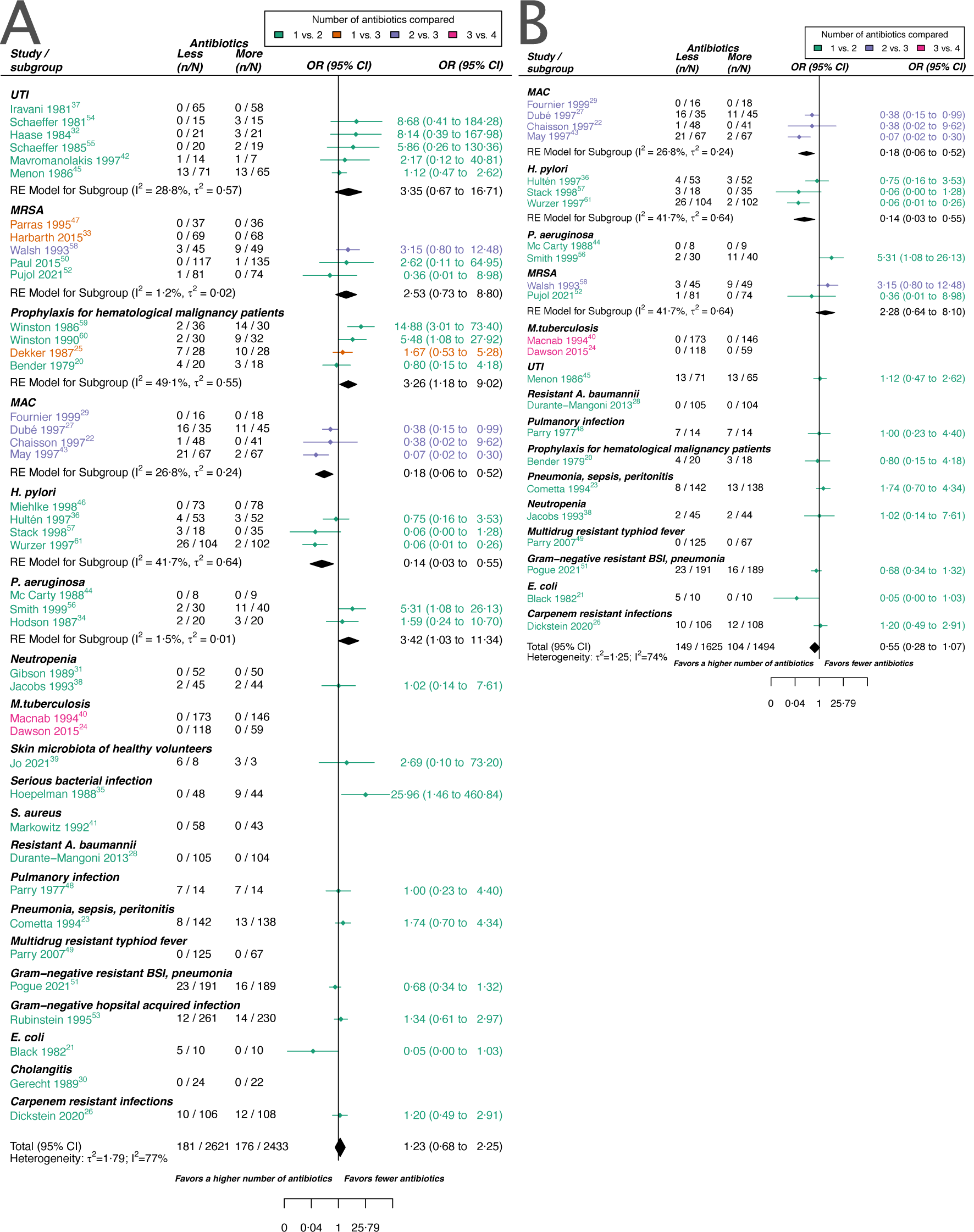
Forest plot of acquisition of bacterial resistance stratified by the reason antibiotics were administered. The coloring indicates the number of antibiotics that were compared in each study. A) The overall pooled LOR of all included studies. B) The pooled LOR of studies with at least one antibiotic in common in the treatment arms. UTI stands for urinary tract infection, MRSA for methicillin-resistant *Staphylococcus aureus*, MAC for *Mycobacterium avium* complex, and BSI for blood stream infection.

The inspection of the funnel plot and the modified Egger’s test showed no indication of a publication bias (SI section 5). The results were largely robust to the choice of the random effects model (SI section 4). The probability of the secondary outcome “alterations of the prescribed treatment due to adverse events”, was higher using more antibiotics in comparison to fewer (pooled OR 1.61, 95% CI 1.12 – 2.31; *I^2^*=5%; SI p 10). In 15 studies (36%), the proportion of patients with alterations of the prescribed treatment due to adverse events was reported, with three studies (20%) reporting zero cases in both treatment arms. All other analyses of secondary outcomes showed no indication of harm or benefit of treating with a higher number of antibiotics (SI section 9).

## Discussion

We performed a meta-analysis of RCTs and quasi-RCTs not limited to a particular bacterial species, specific condition, or antibiotic combinations to assess the effect of antibiotic combination therapy on within-patient resistance development. Our analysis could not identify any benefit or harm of using a higher or a lower number of antibiotics regarding within-patient resistance development. However, we found some evidence that combining antibiotics may be beneficial or harmful for specific pathogens or infection types. Acquisition of resistance was rarely a primary objective of the included RCTs. Hence, they were typically not designed to detect differences in resistance development between treatment arms and underpowered for this endpoint. Therefore, the absence of evidence does not mean that there is convincing evidence for the lack of an effect of using more or fewer antibiotics on resistance development but rather highlights a knowledge gap. This is remarkable given that the general rise of resistance is an increasing concern (3, 18) and a priority area for health policy and public health (62).

Our analysis showed that combining antibiotics reduced resistance development for *H. pylori* or MAC, in line with the current standard of care (11, 63). Surprisingly, we found only two studies that satisfied our inclusion criteria for Mtb (24, 40), which may be considered the prime example of effective antibiotic combination therapy. The limited number of Mtb studies may be because antibiotic administration commonly varies during Mtb treatment, which conflicted with our inclusion criteria that necessitated a consistent treatment regimen for susceptibility measurements (SI section 2). Both eligible Mtb studies were excluded from the analysis due to the absence of any events in either treatment arm.

Our main result, the absence of a general effect of combining antibiotics on resistance development, aligns with the two previous meta-analyses (4, 5). With 42 trials in our systematic review and 29 in the meta-analysis, our study provided a comprehensive assessment of the effect of antibiotic combination therapy on within-patient resistance. Whereas previous meta-analyses focused on a combination of specific antibiotic classes and included fewer than ten studies each, our study aimed to assess the general effect of combining antibiotics on resistance evolution across different bacterial pathogens. By including trials with different antibiotic combinations and bacterial pathogens, we increased clinical and statistical heterogeneity. We accounted for many sources of heterogeneity using stratification and meta-regression, but analyses were limited by missing information and sparse data.

Our findings have implications for the design of future studies of resistance development. Generally, the development of resistance within a patient is a rare event. However, even small differences could be relevant at the population level. To obtain reliable estimates of such differences and to better understand the factors influencing them, very large RCTs would be needed, which systematically investigate the development of antibiotic resistance and include resistance testing of each administered antibiotic. 19 (45%) of our included studies compared treatment arms with no antibiotics in common, and 22 studies (52%) had more than one antibiotic not identical in the treatment arms (table 1). To better evaluate the effect of combination therapy, especially more RCTs would be needed where the basic antibiotic treatment is consistent across both treatment arms, i.e. the antibiotics used in both treatment arms should be identical, except for the additional antibiotic added in the comparator arm (table 1). As such RCTs are costly and associated with high hurdles, the analysis of cohort studies could be an alternative approach. Over 25 years ago, Fish et al. published a systematic summary of prospective observational studies reporting data on resistance development, including antibiotic combination therapy (64). Similarly, today, relevant cohort studies could be analysed collaboratively using various modern statistical methods to address confounding by indication and other biases (65, 66). However, even with appropriate causal inference methods, residual confounding cannot be excluded when using observational data (67). Therefore, RCTs will remain the gold standard to estimate causal relationships.

The main strength of this study is its comprehensive and systematic approach. For one, it allowed identifying a knowledge gap regarding the effect of antibiotic combination therapy on resistance development. Further, our study highlights several issues in the evidence base evaluating antibiotic combination therapy and resistance development. The included trials did not always test and report systematically the susceptibility against all administered antibiotics (table 1). Some antibiotics might have had reduced potency or were ineffective due to pre-existing resistance mutations. Furthermore, in studies where treatment was not targeted against a specific pathogen, some antibiotics may have been inactive against the causative pathogen due to intrinsic resistance. Indeed, one of the reasons for using combination therapy is to broaden the bacterial spectrum for empirical therapy (15), which could contribute to an increased risk of antibiotic resistance spread.

Our study had several limitations. First, despite our systematic search, we might have missed relevant studies. Since resistance development is typically not a primary endpoint and often not reported systematically, relevant trials are challenging to identify. Our search strategy aimed to identify a broad range of trials considering resistance development. However, as a trade-off, our search strategy might have missed trials addressing a specific medical condition or drug combination. Second, our systematic review and meta-analysis included many older studies that did not follow the relevant reporting guidelines (68), thereby hampering data extraction and potentially introducing bias. Third, it is often challenging to discern the specific mechanisms by which resistance develops based on the data from clinical trials. This includes distinguishing whether resistance arises *de novo*, if the pathogen acquires resistance through horizontal gene transfer, if the patient becomes newly infected with a resistant pathogen, or if the pathogen was present but undetected at the beginning of treatment. These scenarios can impact the effectiveness of combination therapy. For example, combination therapy may be more likely to select any pre-existing resistant pathogens compared to monotherapy due to the use of multiple antibiotics. We addressed some of this heterogeneity by employing two different measures of resistance (SI section 1). Furthermore, the variation in standards that classify bacteria as susceptible or resistant adds another layer of heterogeneity alongside the technical limitations in detecting resistance development.

In conclusion, combination therapy offers potential advantages and disadvantages regarding resistance evolution and spread. On the one hand, combination therapy typically increases the genetic barrier to resistance, and it has become the standard therapy for pathogens notorious for resistance evolution. Therefore, combination therapy remains a plausible candidate strategy to slow down resistance evolution. On the other hand, combination therapy generates selection pressure for resistance to multiple antibiotics simultaneously and could, therefore, accelerate resistance evolution – especially in the microbiome. Given the critical nature of this context, it is profoundly disconcerting that there is a lack of evidence elucidating the impact of combining antibiotics on the development of resistance.

## Materials and Methods

### Inclusion criteria and search strategy

We did a systematic review and meta-analysis to summarise the evidence on the effect of antibiotic combination therapy on resistance development. We included RCTs and quasi-RCTs comparing treatments with a higher number of antibiotics to treatments with a lower number of antibiotics. Studies were classified as quasi-RCTs if the allocation of participants to study arms was not truly random. We did not consider antiseptics or compounds supporting the activity of antibiotics, such as beta-lactam inhibitors as antibiotics itself. Whereas the antibiotic substances administered within one treatment arm had to be the same for all patients, the antibiotics could differ between treatment arms. We required baseline and follow-up cultures with resistance measurements to determine the treatment impact on resistance. We considered only antibiotic treatment regimens fixed for the period between two resistance measurements. Hence, we excluded sequential and cycling regimens.

We searched PubMed, EMBASE, and the Cochrane Central Register of Controlled Trials (CENTRAL) from inception up to 24.11.2022, using keywords, medical subject headings (MeSH), and EMTREE terms related to bacterial infection, antibiotics, combination therapy, resistance and RCTs. We excluded complementary and alternative medicine and bismuth. The search strategy is detailed in the SI (section 11). After a systematic deduplication process (69), VNK (or CW) and BS independently screened the titles and abstracts, and, if potentially eligible, the full texts. Any discrepancies between VNK (or CW) and BS were discussed and resolved. At full-text screening, we excluded articles that were not accessible in English or German. We screened the references of eligible studies and the trials included in two previous meta-analyses (4, 5). We followed the PRISMA reporting guidelines (70) and registered our protocol with PROSPERO (CRD42020187257).

### Outcomes

We used two definitions for the primary outcome resistance. A broader definition, “acquisition of resistance”, and a stricter “*de novo* emergence of resistance” definition, where the latter is a subset of the former. A patient was considered to have acquired resistance if, at the follow-up culture, a resistant bacterium (as defined by the study authors) was detected that was not present in the baseline culture. *De novo* emergence of resistance was defined as the detection of a resistant bacterium that was present at baseline but sensitive. Additional secondary outcomes included mortality from all causes and infection, treatment failure overall, treatment failure due to resistance, treatment change due to adverse effects, and acquisition/*de novo* emergence of resistance against non-administered antibiotics. The SI (section 9) provides further details.

### Data extraction and analysis

VNK (or CW) and BS independently extracted all study data using a standardised form (see https://osf.io/gwefy/?view_only=f6a4c1f4c79241038b203bd03c8e1845). The data extracted included the proportion of patients who developed the two primary outcomes and the secondary outcomes and study characteristics such as type of trial (RCT or quasi-RCT), follow-up and treatment duration, number of antibiotics in the treatment arms, type of antibiotic, and presence of comorbidities. Any discrepancies in data extraction were discussed and resolved.

We calculated odds ratios (ORs) with 95% confidence intervals (CIs), comparing a higher with a lower number of antibiotics for each study. We combined ORs using a modified version of the Simmonds and Higgins random effects model (71). If a study had more than two eligible treatment arms, they were merged for statistical analysis. Studies with zero events in both treatment arms were excluded from the statistical analysis. We used subgroup analyses and meta-regressions with multi-model inference to examine the influence of pre-specified variables on summary ORs. Variables included whether the antibiotic(s) used in the arm with the lower number of antibiotics are also part of the arm(s) with the higher number of antibiotics, the number of antibiotics administered, the age of the antibiotics (time since market entry), the administration of other non-antibiotic drugs, whether participants had specific comorbidities or were in intensive care, gram-status of the tested pathogens, and the length of antibiotic treatment and follow-up. We extended our predefined analysis regarding the reason for antibiotic treatment/type of pathogen, which was initially restricted to only *H. pylori* and Mtb, as we found enough studies to stratify by other conditions/pathogens. We furthermore performed post-hoc subgroup analyses to examine the following factors: treatment of resistant pathogens, additional antibiotic administration besides the fixed treatment, and the way of antibiotic administration (SI section 6.2).

Between study heterogeneity was estimated with *I^2^*, using the criteria for *I^2^*specified in Higgins et al. for classifying the degree of heterogeneity (72). CW and BS assessed each study’s quality for the main outcomes using the Risk of Bias tool (RoB 2, SI section 3) (73). To assess publication bias, we visually inspected the funnel plot and a modified Egger’s test (SI section 5). We performed sensitivity analyses on the model choice (SI section 4.1), and risk of bias (SI section 4.2), and performed a post-hoc trial sequential analysis (SI section 8.3). Statistical analyses and visualisations were done in R (version 4.2.1) using packages *metafor* and *MuMIn* (74, 75).

## Data Availability

All data are contained in the the manuscript, supplementary information or online at https://osf.io/gwefy/?view_only=f6a4c1f4c79241038b203bd03c8e1845.

## Acknowledgments

Support from the Swiss National Science Foundation (grant 310030B_176401 (SB, BS, CW), grant 32FP30-174281 (ME), grant 324730_207957 (RDK)) and from the National Institute of Allergy and Infectious Diseases (NIAID, cooperative agreement AI069924 (ME)) is gratefully acknowledged. We thank Anthony Hauser, João Pires and Frédérique Lachmann for helpful discussions and Annelies Zinkernagel and Johannes Nemeth for critical reading of the manuscript. Furthermore, we are grateful for all contacted study authors that responded to our inquiries and provided further information.

