## Supporting Information for "The effect of combining antibiotics on resistance: A systematic review and meta-analysis"

### Supporting Information for
### The effect of combining antibiotics on resistance: A systematic review and meta-analysis

Berit Siedentop[1,2*], Viacheslav N. Kachalov[2,3], Christopher Witzany[1], Matthias Egger[4,5,6], Roger D. Kouyos[2,3†], Sebastian Bonhoeffer[1*†]
[1] Institute of Integrative Biology, Department of Environmental Systems Science, ETH Zürich, Zurich, Switzerland
[2] Division of Infectious Diseases and Hospital Epidemiology, University Hospital Zürich, University of Zürich, Zürich, Switzerland
[3] Institute of Medical Virology, University of Zurich, Zurich, Switzerland
[4] Institute of Social and Preventive Medicine (ISPM), University of Bern, Bern, Switzerland
[5] Population Health Sciences, University of Bristol, Bristol, UK
[6] Centre for Infectious Disease Epidemiology and Research, Faculty of Health Sciences, University of Cape Town, Cape Town, South Africa
[†]These authors contributed equally
*Berit Siedentop, Sebastian Bonhoeffer

**Supporting Information Text**

#### 1. Definitions of resistance development

To measure resistance development in patients with standard clinical routines is challenging. Without antibiotic pressure a resistant strain might be present within the patient at low frequency and might not be detected with a culture due to detection limits. With antibiotic treatment the frequency of this resistant strain might rise and therefore the strain might be detected in a follow-up culture. In this case resistance did not develop *de novo*, but it is difficult to distinguish this case from an event where it did. Furthermore, the genetic relatedness is not always checked between initial and follow-up cultures, meaning that the resistant bacterium at a follow-up culture could have been also transmitted from a different body cite or from other infection sources. To give a more comprehensive overview of how antibiotic treatment strategies might affect the resistance development, we therefore choose to present the results of two resistance estimates. A broader estimate, acquisition of resistance, and a stricter estimate *de novo* emergence of resistance, where the latter is a subset of the former. A patient is considered to have acquired resistance, if at the follow-up culture there has been a resistant (as defined by the study authors) bacterial species detected, that has not been detected in the baseline culture. A patient is considered to have *de novo* emergence of resistance, if at follow-up up culture a resistant bacterium was detected, that has already been detected at the baseline culture, but sensitive. *De novo* emergence of resistance is nested in the definition of acquisition of resistance. In acquisition of resistance we account for bacteria at low abundance that could have been already present at the beginning of treatment, but not detected at screening. In this definition it is impossible to distinguish though, whether the bacteria already colonised the patient or whether the patient was newly infected by an external source during treatment and when the bacterium developed resistance. We also included the stricter definition *de novo* emergence of resistance. For *de novo* emergence we only consider cases where a sensitive bacterium was cultured at baseline. In this definition it is less likely to count cases, where resistant bacteria were transmitted from an external source, as a *de novo* emergence event. But there are cases, which are counted as an event of *de novo* emergence of resistance, where in fact resistance did not develop newly, but resistance was only selected during treatment. This could be the case when a sensitive bacterium was cultured at baseline and the same kind of bacterium was also present at a non-detectable frequency as a resistant phenotype. Overall both resistance development definitions have their limitations and capture slightly different impacts of antibiotic treatment on resistance.

In the main manuscript we showed results for the outcome acquisition of resistance and in the following section the main pooled estimates for *de novo* emergence of resistance are presented.

#### 2. Main estimates for *de novo* emergence of resistance

As for acquisition of resistance (main text figure 3), we did not identify a difference of using a higher number of antibiotics in comparison to less if *de novo* emergence of resistance is considered.

Counterintuitively, for *Mycobacterium tuberculosis* (Mtb) – which may be regarded as the flagship of antibiotic combination therapy – we could only identify two studies matching our inclusion criteria via our systematic search (main text figure 3, figure S1). Since the 1950s the administration of antibiotics often changes within the Mtb treatment period (1, 2). With the early establishment of changing antibiotics within the Mtb treatment period, it would be understandable, that resistance development measurements of periods with fixed antibiotic treatment, which is an inclusion criterion for our review, got less frequent over the years. Therefore, the relatively small proportion of Mtb studies included in our review is not surprising.

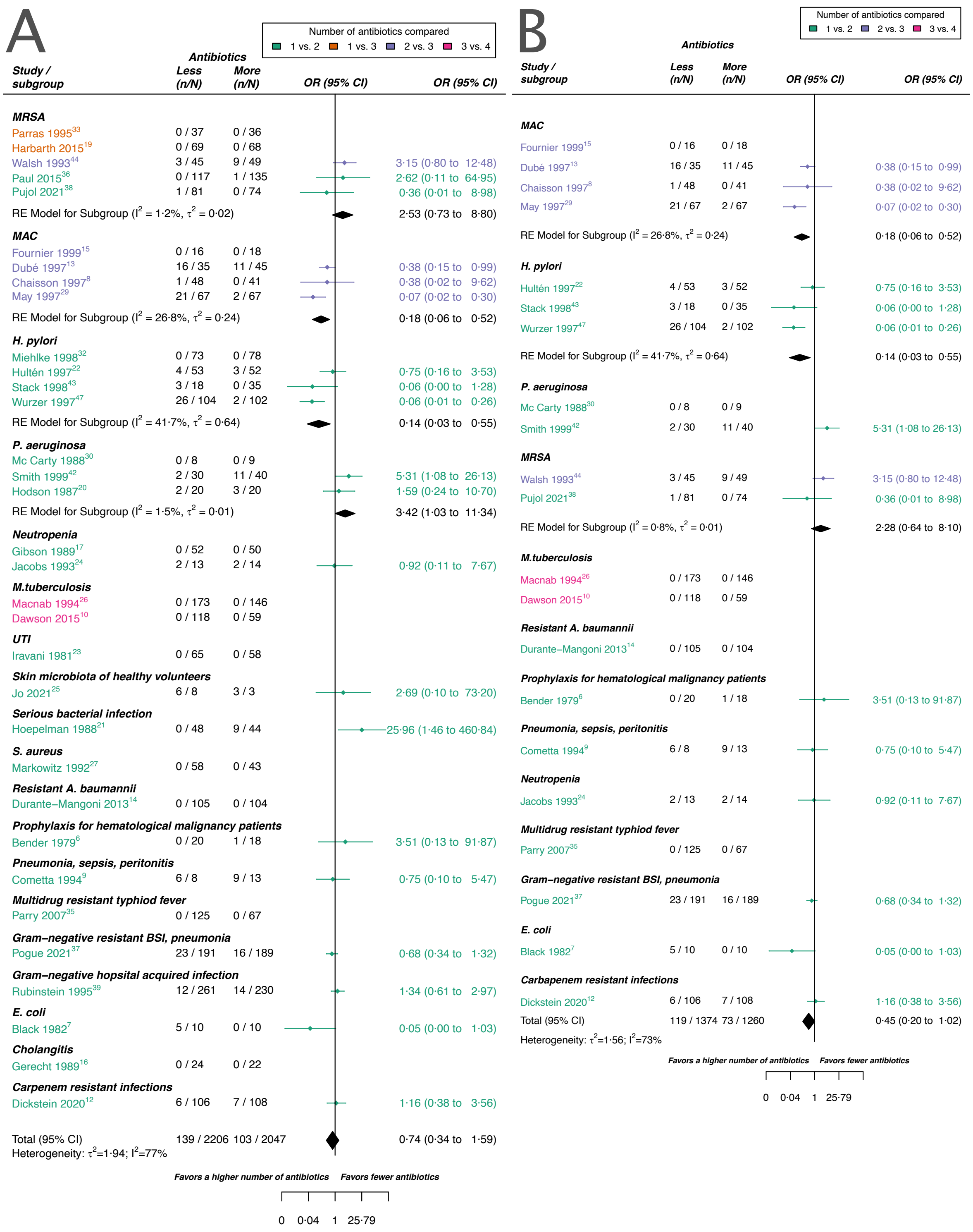


**Fig. S1.** Forest plot of *de novo* emergence of bacterial resistance stratified by the reason antibiotics were administered. The coloring indicates the number of antibiotics that were compared in each study. A) The overall pooled LOR of all included studies. B) The pooled LOR of studies with at least one antibiotic in common in the treatment arms. MRSA stands for methicillin-resistant *Staphylococcus aureus*, MAC for *Mycobacterium avium* complex, and BSI for blood stream infection.

##### *2.1. All studies*

For all studies meeting our inclusion criteria and reporting data of *de novo* emergence of resistance our estimate did not suggest a difference between using a higher number of antibiotics in comparison to less. This result was in line with our main outcome acquisition of resistance. Nevertheless, for *de novo* emergence of resistance there was a slight trend observable which suggested a benefit of using a higher number of antibiotics. However, we could not identify a clear benefit (pooled OR 0.74, 95% CI 0.34 – 1.59, figure S1 A). This trend might be due to the stricter definition of *de novo* emergence relative to acquisition of resistance. In the definition of acquisition of resistance bacterial species that are different from the initial identified infecting organism are included, whereas for *de novo* emergence of resistance they are not necessarily included. For *de novo* emergence of resistance, the efficacy of antibiotic treatment against the considered bacteria is therefore expected to be higher as for acquisition of resistance, as antibiotics typically have a specific bacterial spectrum of activity. The model including all studies reporting *de novo* emergence of resistance showed a substantial amount of heterogeneity ($I^2$=77%, figure S1A).

##### *2.2. Studies with at least one antibiotic common to both treatment arms*

To compare more similar antibiotic treatments, we also estimated the effect of *de novo* emergence of resistance based on studies, that had at least one antibiotic common to the comparator arms. With this restriction we also did not identify a difference of using a higher number of antibiotics in comparison to less, but we observed a stronger tendency of a benefit of using a higher number of antibiotics (pooled OR 0.45, 95% CI 0.20 – 1.02, figure S1B). The model for studies reporting *de novo* emergence of resistance, and with at least one common antibiotic in the comparator arms showed still a substantial amount of heterogeneity ($I^2$=73%, figure S1B).

#### 3. Risk of bias assessment

To assess the risk of bias for our two main outcomes we used the RoB 2 tool (3). The results of the risk of bias assessments for acquisition, and *de novo* emergence of resistance differed only marginally, which can be explained by the overlap of those two definitions. We defined *de novo* emergence of resistance as a stricter subset of acquisition of resistance (section 1). In both cases two studies were classified overall with a low risk of bias, and about 50 % percent of the studies were classified overall with some concerns of bias (67% acquisition of resistance, 72 % emergence of resistance, figure S2). The highest source of at least some concern was the selection of the reported results. As development of resistance is not a typical main objective of RCTs, and since we included a large proportion of rather old studies, the resistance outcome is often not well (pre-)defined (table S1) and not presented in a systematic way, which can explain the risk of bias observed in the category “selection of the reported results”. Since the studies were rather underpowered (main text: figure 2B) to detect the resistance development, missing data was commonly a high risk of concern in the domain “deviations from intended interventions”. The detailed output of the risk of bias assessment using the RoB 2 tool can be found at OSF under the following link “https://osf.io/gwefy/?view_only=f6a4c1f4c79241038b203bd03c8e1845”.

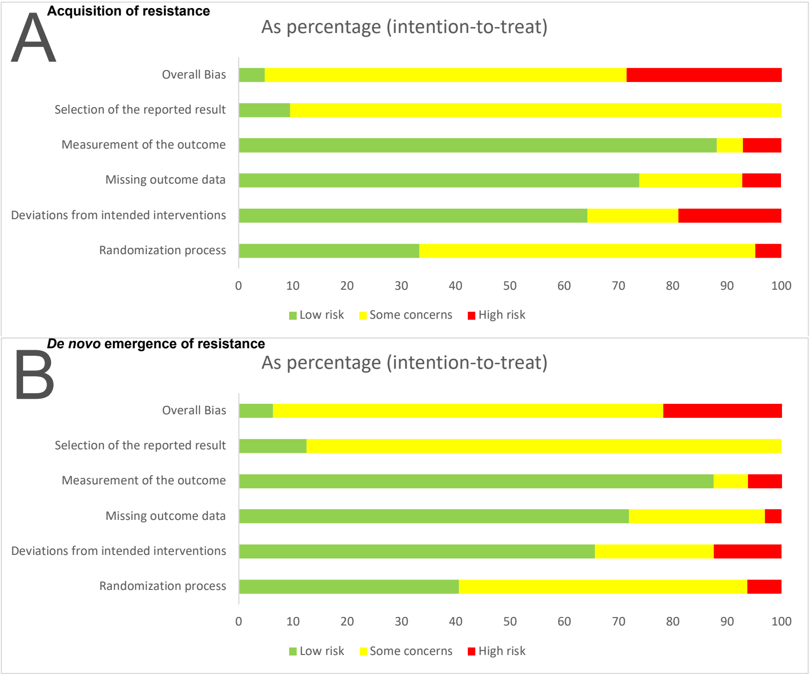


**Fig. S2.** Risk of bias summary for the two main outcomes: A) Acquisition of resistance, B) *de novo* emergence of resistance.

**Table S1.** Justification for extraction of resistance development. The definitions of resistance development are stated as given by study authors. In case no explicit definition was given, we state a justification for extraction and indicate it with (*). Note that for data extraction for the publications of Dekker et al. 2015 and Pogue et al. 2021 additional publications of the same studies were consulted (Paul et al. 2018 (4) and Kaye et al. 2022 (5) respectively). Resistance breakpoints are stated in case numerical values were given in the respective studies. See table 1 in the main text for which antibiotics the studies tested and reported extractable resistance data.

| Study | Definition of resistance development given by study authors or justification for extraction |
|---|---|
| Bender et al. (1979)**(6)** | Susceptibility testing for gentamicin of the flora was performed at randomisation and twice weekly after with Kirby-Bauer disk technique and microtiter minimal inhibitory concentration. (*) |
| Black et al. (1982)**(7)** | Patients were infected with a known strain and all stool cultures and rectal swabs were plated and tested for trimethoprim resistance. (*) |
| Chaisson et al. (1997)**(8)** | Testing of isolates for susceptibility for clarithromycin, ethambutol, and clofazimine was performed before the entry of study and monthly for 6 months in broth by the method of Heifets.(*) |
| Cometta et al. (1994)**(9)** | All microorganisms were sensitive to imipenem at randomisation and follow-up cultures were performed. (*) |
| Dawson et al. (2015)**(10)** | Susceptibility testing at randomisation and for the following cultures by rapid testing. Susceptibilities to isoniazid, rifampicin, and fluoroquinolones were determined by line probe assay. (*) |
| Dekker et al. (1987)**(11)** | At admission cultures were performed and surveillance culture were done twice a week. Gram negative bacilli were tested for antibiotic susceptibility. The minimal inhibitory concentrations were assessed by agar dilution technique. An MIC of ≥ 2 µg/mL was considered resistant for ciprofloxacin, an MIC of ≥ 4 µg/mL for trimethoprim and an MIC ≥ 75 µg/mL for sulfamethoxazole. (*) |
| Dickstein et al. (2019)**(12)** | Development of a new colistin-resistant (ColR) isolate within 28 days from study enrolment. To be considered a new ColR isolate, the ColR isolate had to be detected on Day seven or later in patients for whom the baseline isolate was colistin-susceptible, and for whom no ColR isolate was cultured from the rectal swab taken on Day one. Susceptibility was determined by broth microdilution. Colistin resistance was defined as an MIC > 2mg/L. |

| | |
|---|---|
| Dubé et al. (1997)**(13)** | All available isolates were tested for susceptibility to clarithromycin. Patients were evaluated at the time of enrolment, two and four weeks later, and then every four weeks. Clarithromycin resistance was defined as detectable growth in a concentration of clarithromycin of 8 µg/mL. (*) |
| Durante-Mangoni et al. (2013)**(14)** | The identification of a colistin resistant *Acinetobacter baumannii* during treatment was defined as resistance emergence. Resistance was determined by microdilution method and/or E-test. |
| Fournier et al. (1999)**(15)** | Susceptibility testing was performed at study entry and after 2 months and classification was performed according to Heifets. (*) |
| Gerecht et al. (1989)**(16)** | Emergence of resistance was defined as one cause of treatment failure. Emergence of resistance was classified as the detection of an infecting microorganism resistant to more than 4 µg/mL of gentamicin sulfate or more than 128 µg/mL of mezlocillin sodium during treatment while the patient shows indications of cholangitis. |
| Gibson et al. (1989)**(17)** | Microbiological assessment of the blood was performed before treatment and 96 hours after treatment. (*) |
| Haase et al. (1984)**(18)** | Susceptibility was assessed before therapy, during therapy and after therapy. Susceptibility testing was performed with disk dilution method, and agar dilution method. Resistance results were reported for reinfections defined as the reappearance of infection with a different organism after completion of therapy. Resistance against norfloxacin and trimethoprim-sulfamethoxazole was defined as a larger inhibition zone diameter of 0.17 and 0.16 mm, respectively, or/and a MIC larger than 16 µg/mL and 3.4-64 µg/mL, respectively. (*) |
| Hartbarth et al. (2015)**(19)** | Susceptibility assessment was performed at baseline and at the end of treatment. Susceptibility was performed with a disc diffusion method phenotypically and genotypically. (*) |
| Hodson (1987)**(20)** | *P. aeruginosa* had to be sensitive at inclusion and resistance was measured and reported after 10 days of treatment. Sensitivity was determined by standard disc methods. (*) |
| Hoepelman et al. (1988)**(21)** | Susceptibility was assessed before, during, and after treatment. Susceptibility testing was performed with disc diffusion method and minimum inhibitor concentrations were assessed for blood cultures and patients with no response to treatment with agar dilution technique. Resistance for the agar dilution technique was defined as an MIC of ≥ 32 µg/mL for ceftriaxone, ≥ 8 µg/mL for gentamicin and ≥32 µg/mL for cefuroxime. For the disc diffusion method 30 µg ceftriaxone, 40 µg gentamicin and 60 µg cefuroxime were used. If the zone of inhibition was ≤18 mm cultures were classified as ceftriaxone resistant and sensitive if the zone was ≥26 mm and intermediate in between. For gentamicin the values were ≤20mm and ≥28mm and for cefuroxime ≤20mm and ≥28mm, respectively.(*) |
| Hultén et al. (1997)**(22)** | Susceptibility was assessed by E-test at inclusion and 12 weeks after treatment determination. (*) |
| Iravani et al. (1981)**(23)** | Susceptibility testing at baseline, during treatment and at follow-up. Testing was performed with Bauer's disc diffusion method using 30 µg nalidixic acid, 1.25 µg trimethoprim and 23.75 µg sulfamethoxazole. (*) |
| Jacobs et al. (1993)**(24)** | Emergence of resistance was defined as treatment failure with resistance, i.e., bacteriological failure with the reisolation of original pathogen(s) resistant to the study antibiotic(s) after treatment. |
| Jo et al. (2021)**(25)** | Susceptibility testing before treatment and after treatment by culture. (*) |
| Macnab et al. (1994)**(26)** | Susceptibility testing before treatment and after around 90 doses. (*) |
| Markowitz et al. (1992)**(27)** | Susceptibility was assessed by microdilution method before treatment and for the last continuous positive culture during treatment. Furthermore, susceptibility was assessed for relapse isolates and isolates phenotypically different from the initial one. (*) |
| Mavromanolakis et al. (1997)**(28)** | Susceptibility was assessed before treatment, after 2 weeks, at the end of treatment, and 2 weeks after treatment by disk diffusion method. (*) |
| May et al. (1997)**(29)** | Susceptibility was assessed at treatment start, after two months, and in case of relapse by the Becton Dickinson method. (*) |
| Mc Carty et al. (1988)**(30)** | Susceptibility was assessed at admission, every four days during treatment, and within 48 hours after treatment by broth microdilution method using the American Microscan Gram Negative-Panel. (*) |
| Menon et al. (1986)**(31)** | Susceptibility was assessed before therapy, and after one and two weeks after therapy. (*) |
| Miehlke et al. (1998)**(32)** | Susceptibility was assessed before and after treatment by E-test. An MIC of ≤0.125 mg/L was considered clarithromycin sensitive and an MIC of ≥ 2mg/L resistant. An MIC of ≤2 mg/L was considered amoxicillin susceptible and an MIC of ≥ 4 mg/L resistant. (*) |
| Parras et al. (1995)**(33)** | Susceptibility was assessed at baseline and at end of therapy by agar dilution method or automated microdilution methods. (*) |
| Parry et al. (1977)**(34)** | Susceptibility was assessed before, during, after treatment, after two weeks, and after six months after treatment by Bauer's method. (*) |
| Parry et al. (2007)**(35)** | Susceptibility was assessed before therapy and after treatment by E-test, disk diffusion method. Ofloxacin was tested by disk diffusion method with a 5 µg and organisms were declared susceptible with a breakpoint ≤ 2 µg/mL and resistant with a breakpoint ≥8 µg/mL. Azithromycin |

| | |
|---|---|
| | was also tested with disk diffusion method (15 µg disk), but no clear breakpoint were defined. Instead azithromycin was determined by E-test according to the manufacture's guideline. (*) |
| Paul et al. (2015)**(36)** | Development of resistance was defined as acquisition of *S. aureus* resistant to any of the study drugs or vancomycin resistant *Enterococci*. |
| Pogue et al. (2021)**(37)** | Number of patients, who developed colistin resistance during therapy. Resistance was assessed with broth microdilution and declared as colistin resistant with an MIC ≥4 mg/L. |
| Pujol et al. (2021)**(38)** | Emergence of resistance to study drugs during treatment according to EUCAST. |
| Rubinstein et al. (1995)**(39)** | Resistance emergence was assessed by measuring MICs before, during and after treatment. Disk diffusion testing was performed with disks of 30 µg ceftazidime, 30 µg ceftriaxone and 10 µg tobramycin. An MIC ≤8 mg/L was considered susceptible for ceftazidime and ceftriaxone and a MIC ≥ 32 mg/L was considered resistant for ceftazidime and an MIC ≥ 64 mg/L for ceftriaxone. An MIC ≤4 mg/L was classified as susceptible for tobramycin, and an MIC ≥ 8 mg/L as resistant. |
| Schaeffer et al. (1981)**(40)** | Susceptibility was assessed before therapy, after 7 days, and after five to nine days after therapy by plating. Susceptibility testing was performed by plating 0.1mL of culture on Mac Conkey agar containing 100 µg/mL cinoxacin or 1-24 µg/mL trimethoprim-sulfamethoxazole. Any growing culture was considered resistant and resistance tests were confirmed with standard agar sensitivity testing to a maximum concentration of 100 µg cinoxacin or 80-400 µg trimethoprim-sulfamethoxazole. (*) |
| Schaeffer et al. (1985)**(41)** | Susceptibility testing was performed before therapy, during therapy, and after five to seven days after therapy by plating. 0.1 mL of cultures were plated on either Mueller-Hinton agar containing 10 µg/mL agar of norfloxacin or 1-24 µg/mL agar trimethoprim-sulfamethoxazole with 5 % lysed red blood cells from the horse. Any growing culture was considered resistant and resistance tests were confirmed with tube dilution sensitivity testing to a maximum concentration of 100 µg/mL norfloxacin or 32-608 µg/mL trimethoprim-sulfamethoxazole. (*) |
| Smith et al. (1999)**(42)** | Susceptibility was assessed at inclusion, at end of treatment by disk-susceptibility testing. An MIC of ≥100 µg/mL was considered resistant for azlocillin and resistant to tobramycin if the MIC was ≥8 µg/mL.(*) |
| Stack et al. (1998)**(43)** | Susceptibility was assessed at baseline, and at four or eight weeks after treatment by E-test. Resistance was considered with bacterial growth at a drug concentration of >2 µg/mL for clarithromycin. (*) |
| Walsh et al. (1993)**(44)** | Susceptibility was assessed at baseline and for organisms culturable after the end of therapy and a two-week follow-up period by a microtiter tube dilution technique. Organisms were declared resistant if the MIC was greater than 2 µg/mL for rifampicin, greater than 8 µg/mL for novobiocin, and greater than 2 µg/mL and 38 µg/mL for trimethoprim and sulfamethoxazole. |
| Winston et al. (1986)**(45)** | Susceptibility of surveillance cultures was assessed at baseline, twice weekly during the study period and after study completion. Acquired organisms were defined as new organisms isolated during the study period, that were not present at baseline. An MIC ≤ 16 µg/mL was considered as sensitive for norfloxacin, polymyxin. For disc sensitivity testing cultures were considered sensitive to norfloxacin if a zone of ≥17 mm was present in a 10 µg norfloxacin disk. (*) |
| Winston et al. (1990)**(46)** | New organisms that were isolated during the study period but have not been present before the study were defined as acquired organisms. Susceptibility tests were done by agar dilution method, or by antibiotic disks. An MIC of ≤ 4, 16, or 4 µg/mL for ofloxacin, polymyxin, or vancomycin was considered susceptible to the antibiotics, respectively. For ofloxacin additional disk sensitivity testing was performed. Susceptibility was declared if a zone of 16mm or greater was present around a 5- µg disk of ofloxacin. (*) |
| Wurzer et al. (1997)**(47)** | Susceptibility was assessed pre-treatment and between 4 and 6 weeks of follow-up by agar dilution, and microbroth dilution. An MIC concentration of ≤ 2 µg/mL indicated susceptibility for clarithromycin, and an MIC above 2 µg/mL resistance. A MIC lower or equal to 0.125 µg/mL for amoxycillin was considered susceptible and classified resistant if above 0.125 µg/mL. (*) |

#### 4. Sensitivity analysis for main estimates

To test the robustness of our main analyses, we performed sensitivity analyses based on the model choice and the risk of bias.

##### *4.1. Model choice*

For our analyses we applied the random effects model 4 described in Jackson et al (48) using the R package *metafor (49)*. To test the robustness of our estimates to the model choice we reran the main analyses with the conventional random effects model (model 1 in Jackson et al (48)) and an corresponding Bayesian version of model 4 in Jackson et al (48). For the sensitivity analyses the R packages *metafor* (49), and MetaStan(50) with default settings were used. We observe that our estimates are typically robust to model choice (figures S3, S4). Only for *P. aeruginosa* our estimate was not robust in our sensitivity anlaysis, where the alternative two approaches showed no harm or benefit of using a higher number of antibiotics (figure S3).

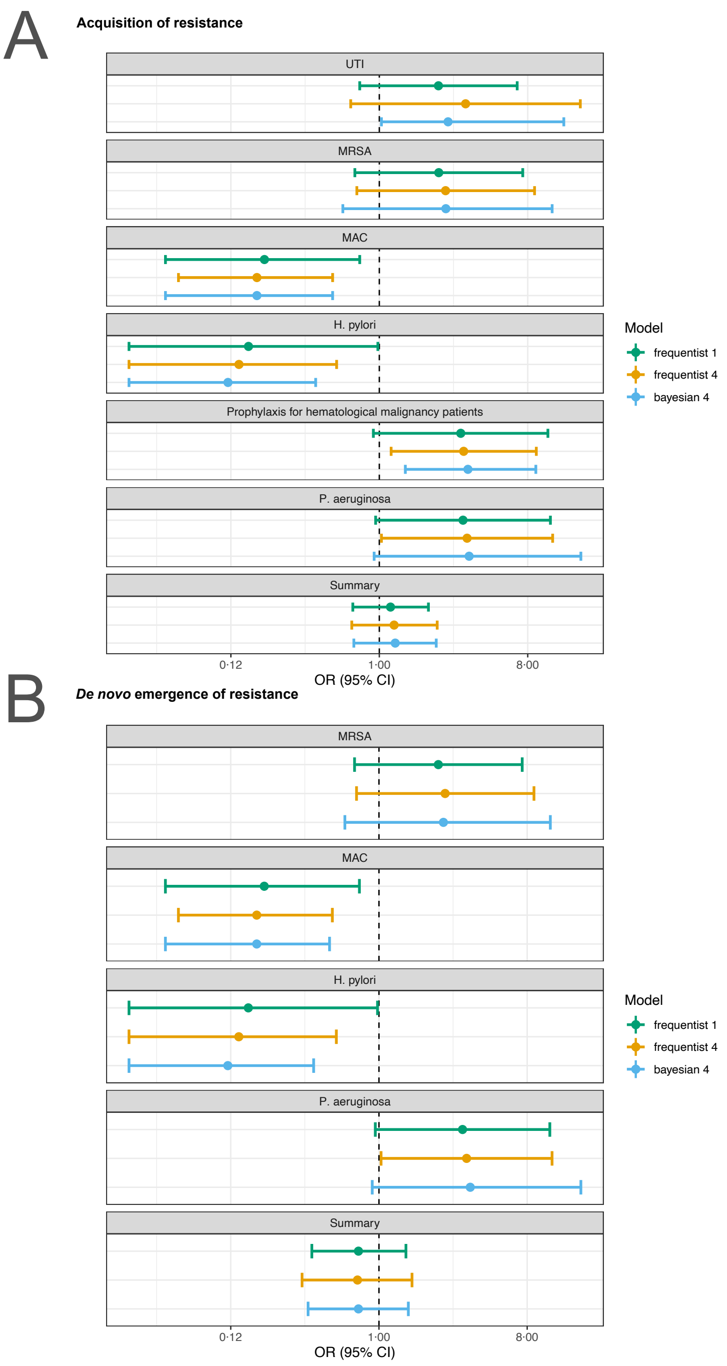


**Fig. S3.** Sensitivity analysis based on model choice for the two main outcomes: A) acquisition of resistance, B) *de novo* emergence of resistance. Shown are the frequentist model estimates of model 1, and model 4 presented in Jackson et al (48) and a Bayesian estimate of model 4. UTI stands for urinary tract infection, MRSA for methicillin-resistant *Staphylococcus aureus*, and MAC for *Mycobacterium avium* complex.

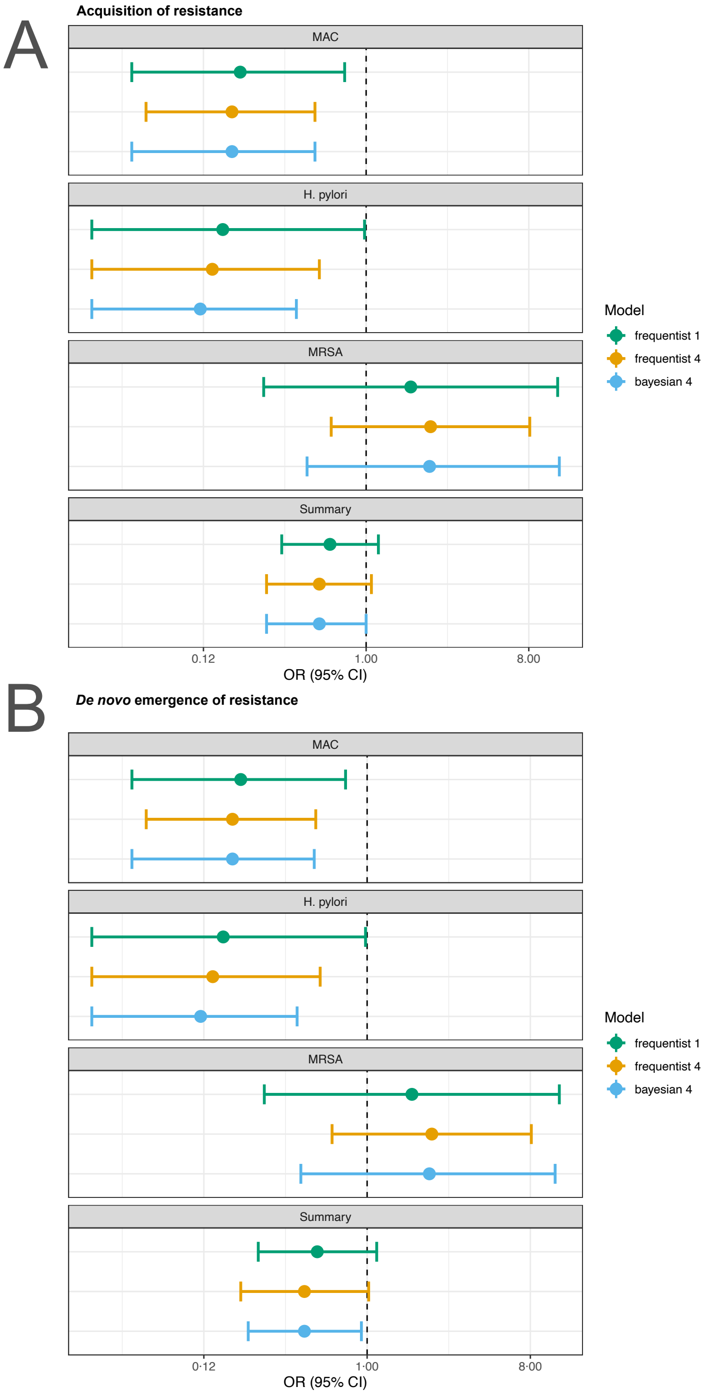


**Fig. S4.** Sensitivity analysis based on model choice for the two main outcomes restricted to studies with at least one common antibiotic in the comparator arms: A) acquisition of resistance, B) *de novo* emergence of resistance. Shown are the frequentist model estimates of model 1, and model 4 presented in Jackson et al (48) and a Bayesian estimate of model 4.

##### *4.2. Impact of risk of bias*

To assess the impact of risk of bias on our estimates, we reran the main analyses stratifying according to the overall risk of bias. For studies classified with an overall high risk of bias our analysis shows that for acquisition of resistance using a lower number of antibiotics shows a benefit (pooled OR 4.45, 95% CI 1.67 – 11.81; $I^2$=57, table S2). We did not observe any difference of using a higher number of antibiotics in comparison to less in resistance development when grouping the rest of the studies according to their risk assessment (table S2). Nevertheless, with less risk of bias administering a higher number of antibiotics seemed to perform better in comparison to less. However, no clear benefit could be determined (table S2). This observation additionally supports that RCTs with resistance development as a main objective, and therefore potentially decreasing the risk of bias, are needed to understand the impact of different treatment strategies on antibiotic resistance outcomes.

**Table S2.** Summary of the results of the sub-group analyses stratifying according to the overall risk of bias for the two main outcomes. Note that the listing of eligible studies also includes studies reporting zero cases in both treatment arms and were therefore not included in the statistical analysis.

| Overall risk of bias | Outcome | OR (95% CI) | Study heterogeneity ($I^2$;$\tau^2$) | Eligible studies |
|---|---|---|---|---|
| Some concerns | Acquisition of resistance | 0.71 (0.38-1.32) | 72%; 1.15 | (6, 7, 9-13, 15, 16, 19, 20, 22, 23, 25, 27-35, 39, 41, 43, 44, 47) |
| Some concerns | *De novo* emergence of resistance | 0.49 (0.21-1.14) | 73%; 1.53 | (6, 7, 9, 10, 12, 13, 15, 16, 19, 20, 22, 23, 25, 27, 29, 30, 32-34, 39, 43, 44, 47) |
| High | Acquisition of resistance | 4.45 (1.67-11.81) | 57%; 1.11 | (8, 17, 18, 21, 24, 26, 36, 37, 40, 42, 45, 46) |
| High | *De novo* emergence of resistance | 2.32 (0.65-8.28) | 60%; 1.28 | (8, 17, 18, 21, 24, 26, 36, 37, 42) |

#### 5. Publication bias

In the study protocol we stated that we will test for publication bias via visual inspection of the funnel plots and by Egger's test. As Egger's test can have problems with false-positive results for dichotomous outcomes, we used a modified version of the Egger's test, i.e. the Harbord's test (51).

Neither the visual inspection of the funnel plots (figure S5), nor Harbord's tests gave an indication for a publication bias for our two main outcomes acquisition, and *de novo* emergence of resistance (acquisition of resistance: Harbord's: p = 0.28; *de novo* emergence of resistance: Harbord's: p = 0.51).

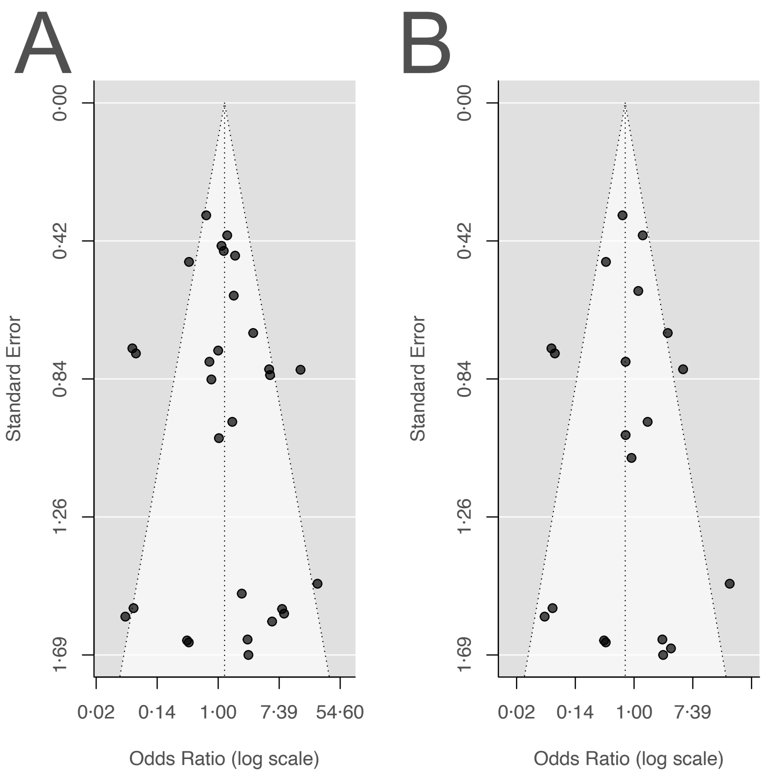


**Fig. S5.** Funnel plots for the two main outcomes: A) acquisition of resistance, B) *de novo* emergence of resistance.

#### 6. Sub-group analyses

The performance of an antibiotic treatment strategy to minimise resistance spread is not only dependent on the number of antibiotics administered. In our main estimates we found a substantial amount of heterogeneity (main text: figure 3; figure S1), which is an indication that additional factors might be important to consider in a statistical model. In the following we first present the results of in the study protocol pre-defined subgroup analyses and afterwards additional post-hoc subgroup analyses. One must consider that the results are mainly based on underpowered studies (main text: figure 2 B), and that in the subgroup analyses the number of included studies decreases. Therefore, the results of the subgroup analyses should be considered with care.

##### *6.1. Predefined in study protocol*

The results of our subgroup-analyses for the outcome acquisition of resistance and de novo emergence of resistance are summarised in table S3 and table S4 respectively. The rationale for carrying out the predefined sub-group analyses are explained in the following subsections.

**Table S3.** Summary of the results of the predefined sub-group analyses for the outcome acquisition of resistance. Note that the listing of eligible studies also includes studies reporting zero cases in both treatment arms, which are not included in the statistical analysis.

| Sub-group Analysis | OR (95% CI) | Study heterogeneity ($I^2$; $\tau^2$) | Eligible studies |
|---|---|---|---|
| **Number of antibiotics administered:** | | | |

| | | | |
|---|---|---|---|
| 1 vs. 2 | 1.49 (0.77-2.88) | 76%; 1.70 | *(6, 7, 9, 12, 14, 16-18, 20-25, 27, 28, 30-32, 34-43, 45-47)* |
| 2 vs. 3 | 0.38 (0.08-1.78) | 74%; 1.63 | (8, 13, 15, 29, 44) |
| **Administration of additional non-antibiotic drugs:** | | | |
| Non-antibiotic drugs as part of treatment | 0.88 (0.21-3.66) | 82%; 3.00 | (6, 11, 22, 32, 33, 43, 45-47) |
| Non-antibiotic drugs administered if necessary | 1.07 (0.48-2.40) | 1%; 0.01 | (12, 14, 23, 24, 38) |
| **Usage of same dosage of antibiotics common to both treatment arms** | 0.59 (0.30-1.18) | 73%; 1.20 | (6, 8, 9, 12-15, 22, 24, 29, 30, 34, 37, 38, 42-44, 47) |
| **Required comorbidity at study inclusion:** | | | |
| Yes | 1.23 (0.50-3.01) | 72%; 1.59 | (6, 8, 11, 13, 15, 17, 20, 24, 27, 29, 30, 34, 42, 45, 46) |
| No | 1.25 (0.55-2.86) | 80%; 2.02 | (7, 9, 10, 12, 14, 16, 18, 19, 21-23, 25, 26, 28, 31-33, 35-41, 43, 44, 47) |
| **Gram-status** | | | |
| Negative | 1.14 (0.56-2.35) | 78%; 1.57 | (7, 11, 12, 14, 20, 22, 23, 25, 28, 30-32, 34, 35, 37, 39, 41-43, 45-47) |
| Positive | 0.44 (0.11-1.76) | 66%; 1.54 | (8, 13, 15, 19, 27, 29, 33, 36, 38, 44) |
| Negative and positive | 3.38 (1.08-10.58) | 44%; 0.75 | (6, 9, 16-18, 21, 24, 40) |
| **Only resistances of antibiotics common to treatment arms** | 0.39 (0.18-0.81) | 75%; 1.49 | (6-10, 12-15, 22, 24, 26, 29-31, 34, 35, 37, 38, 42-44, 47) |
| **Age of antibiotics since conduction of the trial:** | | | |
| Youngest antibiotic is in the treatment arm with the lower number of antibiotics | 1.63 (0.66-4.03) | 76%; 2.17 | (7, 10, 11, 13, 16-22, 24, 30, 31, 33, 38, 41-43, 45, 46) |
| Youngest antibiotic is in the treatment arm with the higher number of antibiotics | 1.08 (0.49-2.42) | 66%; 0.91 | (8, 12, 14, 15, 23, 25, 27-29, 32, 34-37, 39, 40, 44) |
| **No antibiotics common to treatment arms** | 4.73 (2.14-10.42) | 37%; 0.51 | (11, 16-21, 23, 25, 27, 28, 32, 33, 36, 39-41, 45, 46) |

**Table S4.** Summary of the results of the predefined sub-group analyses for the outcome *de novo* emergence of resistance. Note that the listing of eligible studies also includes studies reporting zero cases of resistance in both treatment arms, which were therefore not included in the statistical analysis.

| **Sub-group analysis** | **OR (95% CI)** | **Study heterogeneity ($I^2$; $\tau^2$)** | **Eligible studies** |
|---|---|---|---|
| **Number of antibiotics administered:** | | | |
| 1 vs. 2 | 0.89 (0.38-2.11) | 75%; 1.90 | (6, 7, 9, 12, 14, 16, 17, 20-25, 27, 30, 32, 34, 36-39, 42, 43, 47) |
| 2 vs. 3 | 0.38 (0.08-1.78) | 74%; 1.63 | (8, 13, 15, 29, 44) |
| **Administration of additional non-antibiotic drugs:** | | | |
| Non-antibiotic drugs as part of treatment | 0.22 (0.04-1.10) | 82%; 1.10 | (6, 22, 32, 33, 43, 47) |
| Non-antibiotic drugs administered if necessary | 0.97 (0.36-2.58) | 1%; 0.01 | (12, 14, 23, 24, 38) |
| **Usage of same dosage of antibiotics common to both treatment arms** | 0.53 (0.24-1.16) | 71%; 1.38 | (6, 8, 9, 12-15, 22, 24, 29, 30, 34, 37, 38, 42-44, 47) |
| **Required comorbidity at study inclusion:** | | | |
| Yes | 0.71 (0.21-2.41) | 67%; 1.57 | (6-10, 12-15, 22, 24, 26, 29, 30, 34, 37, 38, 42-44, 47) |
| No | 0.75 (0.28-2.01) | 80%; 2.02 | (16, 17, 19-21, 23, 25, 27, 32, 33, 36, 39) |

| | | | |
|---|---|---|---|
| **Gram status** | | | |
| Negative | 0.60 (0.23-1.55) | 78%; 1.59 | (7, 12, 14, 20, 22, 23, 25, 30, 32, 34, 37, 39, 42, 43, 47) |
| Positive | 0.44 (0.11-1.76) | 66%; 1.54 | (8, 13, 15, 19, 27, 29, 33, 36, 38, 44) |
| Negative and positive | 3.34 (0.59-18.97) | 47%; 1.39 | (6, 9, 16, 17, 21, 24) |
| **Only resistances of antibiotics common to treatment arms** | 0.32 (0.16-0.66) | 59%; 0.87 | (6-10, 12-15, 22, 24, 26, 29, 30, 34, 37, 38, 42-44, 47) |
| **Age of antibiotics since conduction of the trial:** | | | |
| Youngest antibiotic is in the treatment arm with the lower number of antibiotics | 0.73 (0.19-2.77) | 75%; 2.83 | (7, 10, 13, 16, 17, 19-22, 24, 30, 33, 38, 42, 43) |
| Youngest antibiotic is in the treatment arm with the higher number of antibiotics | 0.86 (0.34-2.17) | 70%; 0.98 | (8, 12, 14, 15, 23, 25, 27, 29, 32, 34, 36, 37, 39, 44) |
| **No antibiotics common to treatment arms** | 3.54 (0.91-13.75) | 38%; 0.68 | (16, 17, 19-21, 23, 25, 27, 32, 33, 36, 39) |

###### 6.1.1. Number of antibiotics administered

In our systematic review we did not predefine a fixed number of antibiotics to compare. We rather aimed to investigate whether there is a general trend of a treatment strategy with a higher number of antibiotics performing better than one with less antibiotics with respect to resistance development. One can imagine though, that the magnitude of this trend might vary depending on the number of antibiotics compared. For example, if resistance against the used antibiotics is likely to be encountered in the population, a comparison of one versus two antibiotics might give different results than two versus three. In the 1960s for Mtb the number of antibiotics was for instance increased to three antibiotics at the initial treatment phase, due to the finding that primary resistance can be encountered for one drug but rarely to two or three antibiotics (2, 52, 53). On one hand, if the number of antibiotics used is rather high in both treatment arms, there might be no difference in resistance development detected as the treatment period might be too short to observe a relevant effect. On the other hand, if the treatment period is rather long, there might also not be an efficient effect detectable when a low number of antibiotics is compared and the timespan between follow-up cultures is long. We considered the effect of treatment length, and length of follow-up on our estimates later in the meta-regression and multi-model inference (section 7).

We identified three studies comparing one versus three antibiotics, but two of them had zero events for both comparator arms. We included two Mtb studies in our review comparing three versus four antibiotics, but both had zero events in the comparator arms. For the estimates one versus two antibiotics and two versus three antibiotics we did not identify a difference of using a higher number of antibiotics in comparison to less, and substantial heterogeneity was observed (tables S3, S4). Nevertheless, for the estimate two versus three antibiotics there was a beneficial trend for using a higher number of antibiotics observable. However, no clear benefit could be determined (tables S3, S4). This might indicate that in general a higher number of antibiotics in treatments is beneficial.

###### 6.1.2. Administration of additional non-antibiotic drugs

In our inclusion criteria we allowed the administration of additional non-antibiotic drugs, which potentially could also affect the resistance outcome due to faster cure of patients, or by specifically supporting the activity of antibiotics, as e.g. beta-lactam-inhibitors. To test the effect of the administration of additional non-administered antibiotics, we performed a sub-group analysis based on whether a study administered additional non-antibiotic drugs or not. Notably, in our studies the additional non-antibiotics were always administered in both treatment arms. Considering if additional non-antibiotic drugs were administered or not did not show any harm or benefit on the resistance outcome whether a higher number of antibiotics was used or a lower number (tables S3, S4). A few studies allowed the administration of additional non-antibiotics, but they were not a fixed part of the treatment regime. Also, in those studies we did not identify a harm or benefit (tables S3, S4).

###### 6.1.3. Usage of same dosage of antibiotics common to both treatment arms

Not only the number of total antibiotics might determine the efficacy of a treatment, but also the dosage of antibiotics. To compare more similar treatments, we estimated the pooled OR for studies that administered at least one antibiotic common to both treatment arms, and where additionally the antibiotics that were common were administered with the same dosage. We observed that in most cases if at least one common antibiotic was administered, their dosage was the same (78% acquisition of resistance, 86% emergence of resistance). Therefore, it is not surprising that we observe, in line with the analysis “at least one antibiotic common to both treatment arms” (main text: figure 3B, figure S1 B), no difference in using a higher number of antibiotics in comparison to less to reduce resistances (tables S3, S4). In both cases we observed a substantial amount of heterogeneity, which indicates that further factors might play a role for explaining the observed resistance differences.

###### 6.1.4. Required comorbidity at study inclusion

The way the immune-system reacts to an infection might potentially influence the frequencies of resistances observed (54). Therefore, we tested whether studies that considered patients with a comorbidity, assuming that the immune system is to some extent compromised, show a different trend of resistance development in comparison to studies where no comorbidity was required for study inclusion. For this analysis we considered studies, that had comorbidities as a requirement for study entry. We could not identify a difference of using a higher number of antibiotics in comparison to less for both main outcomes, and regardless of comorbidity status at study entrance (tables S3, S4).

###### 6.1.5. Study was conducted in an ICU

Another way to test the potential role of the immune system is by severity of illness, approximated whether the study population was treated within an ICU or not. We were not able to link on a patient level the data of resistance development to the patient’s ICU status. Therefore, we tried to classify the ICU status per study, i.e. one status for the whole study population. We only identified two studies (5%) for acquisition of resistance, where the whole study population was in the ICU. We found 9 studies (21%), where no patient was treated in the ICU. For the rest of the studies the population could either be mixed (14%), or no information was confidentially extractable (60%). Since the ICU status on a study level seemed to be an uninformative proxy, we decided not to perform sub-group analyses for this factor.

###### 6.1.6. Gram-status

The gram status of a bacterium may potentially determine how effective an antibiotic, or an antibiotic combination is. Differences between gram-negative and gram-positive bacteria such as distinct bacterial surface organisation can lead to specific intrinsic resistances of gram-negative and gram-positive bacteria against antibiotics (55). These structural differences can lead to varying effects of antibiotic combinations between gram-negative and gram-positive bacteria (56). Additionally, plasmids play a major role in the dissemination of antibiotic resistance genes in both gram-positive, and negative bacteria (57). The spread of plasmids differs considerably between gram-positive bacteria and gram-negative bacteria (58). These structural differences could influence the performance of antibiotic treatment strategies. To test the influence of the gram-status on our estimates we performed sub-group analyses with studies, that focused only on measurements of gram-negative bacteria, gram-positive, or both. We classified the gram-status on a study level as we could not link the gram-status and resistance development on a patient level.

When selecting for studies that either focus on gram-negative, or gram-positive we did not identify a difference in using a higher number of antibiotics in comparison to less for both main outcomes (tables S3, S4). For the subgroup analysis including studies with the focus on both gram-negative and positive bacteria the treatment strategy with a lower number of antibiotics showed a benefit for the main outcome acquisition of resistance (pooled OR 3.38, 95% CI 1.08 – 10.58; $I^2$=44%, table S3). However, for *de novo* emergence of resistance we did not identify a difference (pooled OR 3.35, 95% CI 0.67 – 16.71; $I^2$=47%, table S4) It seems, that acquisition of resistance is more sensible to the restriction on which gram-status is considered. This might be due to the broader definition of acquisition of resistance as it is more sensitive to resistance changes in the microbial community. If a treatment is targeted against a specific pathogen, e.g. a gram-positive bacterium, other bacteria of the microbiota are exposed to the treatment as well. Some bacteria of the microbiota might be more intrinsically resistant against the administered antibiotics, e.g. a gram-negative bacterium, and are therefore more likely to develop resistance. With acquisition of resistance, we might detect such effects.

###### 6.1.7. All resistances not only against administered antibiotics

Antibiotic resistances can be acquired by plasmids, which in a clinical context often confer resistances against multiple antibiotics (59-61). Therefore, we aimed to test, whether a higher number of antibiotics also leads to resistance against a higher number of antibiotics, considering both resistances of antibiotics that were administered and ones that were not. For acquisition of resistance, we only identified seven studies that measured resistances also against non-administered drugs. Only three of those studies have non-zero events. For *de novo* emergence of resistance, we identified four studies measuring resistances against non-administered antibiotics, were two of them have non-zero events in both treatment arms. Due to the small number of studies identified and even smaller number of studies having non-zero resistance events, we only present the estimates of the resistances against non-administered antibiotics (sections 9.6 and 9.7).

###### 6.1.8. Only resistances of antibiotics common to treatments arms

To estimate how the same antibiotics performed in the different treatment arms we performed a subgroup-analysis only considering resistance against antibiotics common to both treatment arms. For both main outcomes we observed that if we only consider resistances of common antibiotics the treatment arm with the higher number of antibiotics showed a benefit (acquisition of resistance: pooled OR 0.39, 95% CI 0.18 – 0.81; $I^2$=76%, table S3; emergence of resistance: pooled OR 0.32, 95% CI 0.16 – 0.66; $I^2$=59% table S4). Consequently, we can conclude that for a specific antibiotic less resistances will develop in a treatment arm with a higher number of antibiotics.

As the studies included in our meta-analysis often did not quantify the resistance outcome for all antibiotics administered in a treatment arm it is harder to assess the full resistance burden of the antibiotic treatments systematically. One could argue that due to the higher number of antibiotics given in one treatment arm, one would also observe in total a higher resistance burden in that arm. This possible effect could be magnified dependent on the potency of antibiotics. If a treatment arm is a combination of a low potency antibiotics, one might expect a higher chance of resistance. The results of this sub-group analysis highlight once more that a systematic exploration of resistance development in RCTs is important for a better understanding of resistance development during treatment and that the identity of the administered antibiotics might play an important role.

###### 6.1.9. Age of antibiotics since conduction of the trial

The prevalence of antibiotic resistance affects the treatment success. If resistance before treatment is frequent in the population, then this increases the likelihood that the prescribed antibiotic treatment fails for any patient. We collected data on the year the admission of patients for the individual studies started and the year antibiotics became available. With the naive assumption that the longer the antibiotic has been available before the study was conducted, the higher is its resistance prevalence within the population. This assumption has its weaknesses as antibiotics are used with different intensities over the years and their local pattern of use might vary. However, such data are more difficult to retrieve. Hence, the years an antibiotic was available until the trial started is a simple first approximation to investigate resistance prevalence.

If the studies did not state the year the trial started, we extracted the publication year. For the availability of antibiotics, we used the older of the two dates available on DrugBank (62) and DrugCentral (63) (DrugBank: marketing start , DrugCentral: approvals).

In the following we present the subgroup analyses, where we classify in which comparator arm the youngest antibiotic is administered. We did not detect a harm or benefit of using a higher or lower number of antibiotics when stratifying, and observed in all subgroup analyses at least a substantial amount of heterogeneity (table S3, table S4).

Furthermore, we performed subgroup analyses stratifying according to the mean age of antibiotics in a treatment arm, and the oldest antibiotic of the treatment arm. For those analyses we also did not identify a difference of using a higher number of antibiotics over fewer. It could be that our approximation is too simplified to estimate the potential effect.

###### 6.1.10. No antibiotics common to treatment arms

In the main analyses we presented the estimates for all studies, and studies, which administered at least one antibiotic common to the treatment arms. Here in the supplement we present the resistance estimates for less comparable treatments, i.e. for studies, whose treatment arms had no antibiotics in common. For those studies we observed for both main outcomes a trend favouring the treatment arm with fewer antibiotics (acquisition of resistance: pooled OR 4.73, 95% CI 2.14 – 10.42; $I^2$=37%, table S3; *de novo* emergence of resistance: pooled OR 3.54, 95% CI 0.91 – 13.75; $I^2$=38%, table S4). The benefit was for acquisition of resistance clear, and for *de novo* emergence of resistance not. The result that if the treatment arms had no antibiotics in common a lower number of antibiotics performed better than a higher number of antibiotics could be due to different potencies of antibiotics or resistance prevalences. Further, there could be a bias to combine less potent antibiotics or antibiotics with higher resistance prevalence to ensure treatment efficacy, which could lead to higher chances to detect resistances in the treatment arm with higher number of antibiotics, e.g. by selecting pre-existing resistance (see also section 6.1.9). This highlights once more that the identity of antibiotics may play an important role in determining whether combining antibiotics is beneficial or not with respect to resistance development.

###### 6.1.11. Systematic testing of the whole study population

In our protocol we predefined that we would perform a sub-group analyses based on whether the resistance data were systematically available for the whole study population or just a subset of patients. All our included studies attempted to measure resistance data for the whole study population. In some cases, more information on resistance development was reported than what we could use. In those cases, it was impossible to distinguish how many patients were evaluable for the resistance outcomes, and/or how many patients developed resistances. In summary, we always obtained data for the whole study population, except of the missing data cases, but nevertheless we could not process all information given due to the way it was reported. The influence of missing data is assessed in the risk of bias assessment (section 3), and the corresponding sensitivity analyses (section 4.2).

##### 6.2. Post-hoc subgroup analyses

###### 6.2.1. Additional administration of antibiotics

During our selection process of studies, we realised that some studies allowed the addition of further antibiotics to the assigned treatments, if necessary, whereas others explicitly stated no other antibiotics than the assigned ones are given during the treatment phase. For a large proportion of all included studies, we could not extract whether additional antibiotics were allowed or not (62%). As we cannot rule out that in those studies no additional antibiotics were administered, we decided to include studies where additional antibiotics are allowed. To check the impact of this decision we performed a sub-group analyses for those studies, where information of administration of additional antibiotics was given. We identified 12 studies, which allowed the administration of additional antibiotics, but only at most seven studies could be included in the statistical analyses as the other trials reported zero cases in both treatment arms (14, 17, 19, 23, 30) (tables S5, S6). We identified three studies explicitly excluding additional antibiotics, however the statistical analyses is based on two studies as one reported zero cases in both treatment arms (16) (tables S5, S6). Therefore, the impact of allowing the administration of additional antibiotics, if necessary, on our overall estimates was difficult to infer.

**Table S5.** Summary of the results of the post-hoc sub-group analyses for the outcome acquisition of resistance. Note that the listing of eligible studies also includes studies reporting zero cases in both treatment arms and were therefore not included in the statistical analysis.

| Sub-group Analysis | OR (95% CI) | Study heterogeneity ($I^2$; $\tau^2$) | Eligible studies |
|---|---|---|---|
| **Additional administration of antibiotics:** | | | |
| Allowed | 1.18 (0.70-1.97) | 16%; 0.07 | *(12, 14, 17, 19, 23, 24, 30, 36-39, 46)* |
| Prohibited | 0.19 (0.04-0.98) | 57%; 0.79 | (16, 22, 47) |
| **Pre-resistance against non-administered** | | | |

| | | | |
|---|---|---|---|
| **antibiotics required at study inclusion:** | | | |
| Required | 1.08 (0.57-2.05) | 15%; 0.07 | (12, 14, 19, 33, 36-38, 44) |
| No | 1.25 (0.61-2.55) | 79%; 2.22 | (6-11, 13, 15-18, 20-32, 34, 35, 39-43, 45-47) |
| **Way of antibiotic administration:** | | | |
| Orally | 1.18 (0.44-3.15) | 78%; 2.70 | (6-8, 10, 13, 18, 22, 23, 25, 26, 28, 31, 32, 35, 40, 41, 43-47) |
| Intravenously | 1.83 (0.67-5.00) | 66%; 0.90 | (12, 14, 16, 17, 21, 24, 27, 37, 38, 42) |
| Different ways of administration in the treatment arms | 1.51 (0.67-3.39) | 1%; 0.01 | (11, 20, 34, 36) |

**Table S6.** Summary of the results of the post-hoc sub-group analyses for the outcome *de novo* emergence of resistance. Note that the listing of eligible studies also includes studies reporting zero cases in both treatment arms and were therefore not included in the statistical analysis.

| Sub-group Analysis | OR (95% CI) | Study heterogeneity ($I^2$; $\tau^2$) | Eligible studies |
|---|---|---|---|
| **Additional administration of antibiotics:** | | | |
| Allowed | 0.95 (0.59-1.51) | 3%; 0.01 | *(12, 14, 17, 19, 23, 24, 30, 36-39)* |
| Prohibited | 0.19 (0.04-0.98) | 57%; 0.79 | (16, 22, 47) |
| **Pre-resistance against non-administered antibiotics required at study inclusion:** | | | |
| Required | 1.07 (0.53-2.18) | 17%; 0.10 | (12, 14, 19, 33, 36-38, 44) |
| No | 0.63 (0.23-1.68) | 78%; 2.57 | (6-10, 13, 15-17, 20-27, 29, 30, 32, 34, 39, 42, 43, 47) |
| **Way of antibiotic administration:** | | | |
| Orally | 0.37 (0.11-1.23) | 69%; 1.96 | (6-8, 10, 13, 22, 23, 25, 26, 32, 43, 44, 47) |
| Intravenously | 1.82 (0.64-5.18) | 66%; 0.90 | (12, 14, 16, 17, 21, 24, 27, 37, 38, 42) |
| Different ways of administration in the treatment arms | 2.12 (0.35-12.79) | 1%; 0.01 | (20, 34, 36) |

###### 6.2.2. Pre-resistance against non-administered antibiotics

Some of the studies we included were focused on the treatment of resistant pathogens. Therefore, we tested whether carriage of resistance against non-administered antibiotics might affect the development of resistance against administered antibiotics. We identified eight studies requiring pre-resistance, of which five had non-zero events in both treatment arms. For both studies requiring pre-resistance and no pre-resistance, we could not identify a trend favouring more or less antibiotics (tables S5, S6). As multi-drug resistance is an increasing concern it is important to understand if the optimal treatment strategy for pre-resistant pathogens might differ from the one of sensitive pathogens. However, the data of our meta-analysis are not sufficient to answer this question.

###### 6.2.3. Way of antibiotic administration

The way how antibiotics are administered, e.g. intravenously (IV) or orally, could also impact the development of antibiotic resistance due to different pharmacokinetics and potential differing antibiotic bioavailability (64). Therefore, we stratified our studies according to the way antibiotics were administered: orally, or IV in both treatment arms, or the way of administration differed in the treatment arms. We could not identify a harm or benefit in the sub-group analyses of using a higher or a lower number of antibiotics (tables S5, S6).

#### 7. Meta-regressions and multi-model inference

Additionally, to the subgroup analyses we also performed meta-regressions for the exploration of the importance of factors potentially affecting our main outcomes. For the meta-regression

models we used the conventional random effects model (model 1 in Jackson et al (48)) due to convergence issues with model 4 and since our sensitivity analysis of the main outcomes showed typically robustness to the model choice (section 4). With performing meta-regressions, we were able to include continuous covariables such as treatment length, and by multi model inference we could obtain parameter estimates averaged over a set of models. The set of possible models was restricted to meta-regression models with up to two covariables and no interaction terms to avoid overfitting. We performed multi-model inference with the R package *MuMIn* (version 1.46) (65).
For the multi-model inference all meta regression models of the set of possible models were simulated. Following a model selection approach using the Akaike information criterion (AIC) model, the AIC value for each model was calculated. The AIC is a measure of fit, which is based on the log-likelihood function, and the number of unknown model parameters. Smaller AIC values are assigned to better model fits. In addition to the AIC value, we calculated the AIC differences (ΔAIC) between each model and the model with the lowest AIC value. With ΔAIC we calculated the Akaike weights, which can be interpreted as the probability a model is the best of the given set of models and data. With the full model approach, we then calculated the model averaged coefficients, which are estimates weighted by the Akaike weights and averaged over the whole set of possible models (full model average). For the interested reader further, detailed information can be found in literature about multi-model inference (66-68).
The covariables we considered for the meta-regression included: (i) administration of antibiotics common to the treatment arms, (ii) required comorbidity status at study inclusion, (iii) the year difference between the youngest antibiotic in the treatment arm with a lower number of antibiotics and the youngest in the treatment arm with a higher number of antibiotics, (iv) the treatment length, (v) the length of study/resistance follow up, (vi) gram status of bacteria with resistance measurements, and (vii) the number of antibiotics administered.
In some cases, the treatment length of the two treatment arms within a study were of different length, in those cases we took as the treatment length covariable the average treatment time of both treatment arms. As the treatment times between treatment arms did not vary a lot, we did not explore those differences further. Furthermore, we wanted to consider the age of antibiotics since the conduction of a trial. There are several ways of how to implement this as a covariable. We decided to take the difference of the youngest antibiotics in both treatment arms, as we expected that novel antibiotics are more likely to be tested in the treatment arm with lower antibiotics.
For our multi-model inference, we excluded the variables considering whether a study was conducted in an ICU, and whether additional drugs were administered as we could not confidently obtain information regarding those variables for more than half of the studies. Due to a high correlation between the administration of antibiotics common to the treatment arms and same dosage (acquisition of resistance: 0.95, *de novo* emergence of resistance: 0.91) we excluded the variable same dosage from the meta-regressions.
Our multi model inference showed that for acquisition of resistance the most important covariable to include in a meta-regression model to explain some of the observed heterogeneity was whether antibiotics common to the treatments were used or not (table S7). This is in line with our sub-group analysis performed (main text figure 3B). By including the information whether at least one antibiotic was common to both treatment arms in a meta-regression, we could find a decrease in the estimated heterogeneity ($I^2$=59, no-meta-regression: $I^2$=77), but nevertheless the heterogeneity remains substantial (table S8). Furthermore, we could confirm once more that a lower number of antibiotics performs better, if in the treatment arms no common antibiotics are used (tables S7, S8). For *de novo* emergence of resistance, the multi model inference did not show any significant covariables (table S9).
Overall, this does not necessarily mean that any of the covariables are not impacting the outcome of resistance development significantly, but since most studies were underpowered (main text figure 2 B) there is the possibility that we are missing important signals.

**Table S7.** Overview of the model averaged coefficients obtained by the multi-model inference for the main outcome acquisition of resistance. Significant model estimates are displayed in a bold font.

| Model-averaged coefficients (full-average) | Estimated | Standard error | z value | Pr(>\|z\|) |
|---|---|---|---|---|
| **Intercept** | 0.73 | 1.69 | 0.60 | 0.54 |
| **Length of follow-up** | 1.00 | 1.00 | 0.46 | 0.65 |
| **Treatment length** | 0.99 | 0.01 | 0.77 | 0.43 |
| **1 vs. 3 antibiotics** | 0.78 | 2.19 | 0.31 | 0.75 |
| **2 vs. 3 antibiotics** | 1.16 | 2.16 | 0.19 | 0.85 |
| **Antibiotics in common: no** | **5.67** | **1.89** | **2.73** | **0.01** |
| **Comorbidity: yes** | 1.35 | 1.77 | 0.53 | 0.60 |
| **Gram positive and negative bacteria** | 1.52 | 1.91 | 0.65 | 0.52 |
| **Gram positive bacteria** | 1.08 | 2.07 | 0.11 | 0.91 |
| **Year difference of youngest antibiotics** | 1.00 | 1.01 | 0.15 | 0.88 |

**Table S8.** Model output for a meta-regression for acquisition of resistance including as a covariable, whether at least one antibiotic was in common in the treatment arms. Significant model estimates are displayed in a bold font.

| | OR (95% CI) | z value | Pr(>\|z\|) | Study heterogeneity ($I^2$; $\tau^2$) |
|---|---|---|---|---|
| **Intercept** | 0.63 (0.33-1.21) | -1.39 | 0.17 | 59%; 0.90 |
| **Antibiotics common: no** | 5.86 (2.05-16.76) | 3.30 | <0.01 | |

**Table S9.** Overview of the model averaged coefficients obtained by the multi-model inference for the main outcome *de novo* emergence of resistance.

| Model-averaged coefficients (full-average) | Estimated | Standard error | z value | Pr(>\|z\|) |
|---|---|---|---|---|
| **Intercept** | 2.22 | 2.42 | 0.90 | 0.37 |
| **Length of follow-up** | 0.99 | 1.01 | 0.73 | 0.46 |
| **Treatment length** | 1.00 | 1.01 | 0.41 | 0.68 |
| **2 vs. 3 antibiotics** | 1.29 | 2.51 | 0.28 | 0.78 |
| **Antibiotics in common: yes** | 0.32 | 2.66 | 1.16 | 0.25 |
| **Comorbidity: yes** | 1.40 | 2.03 | 0.47 | 0.64 |
| **Gram positive and negative bacteria** | 1.45 | 2.23 | 0.47 | 0.64 |
| **Gram positive bacteria** | 1.16 | 1.98 | 0.21 | 0.83 |
| **Year difference of youngest antibiotics** | 0.99 | 1.02 | 0.30 | 0.72 |

#### 8. Statistical power

##### *8.1. Adequate treatment arm size*

Resistance development is a rare event and therefore differences in resistance development are difficult to detect in small population sizes. To illustrate this, we calculated how much participants would have needed to be included per treatment harm in order to detect whether a higher number of antibiotics would half the odds of occurrence of resistance and compared it to the actual number of participants (figure S6). For the calculations we assumed a power of 80% and used for each trial the upper confidence interval for the probability of resistance development in the treatment arm with the lower number of antibiotics. The confidence interval was determined with Bayesian inference.

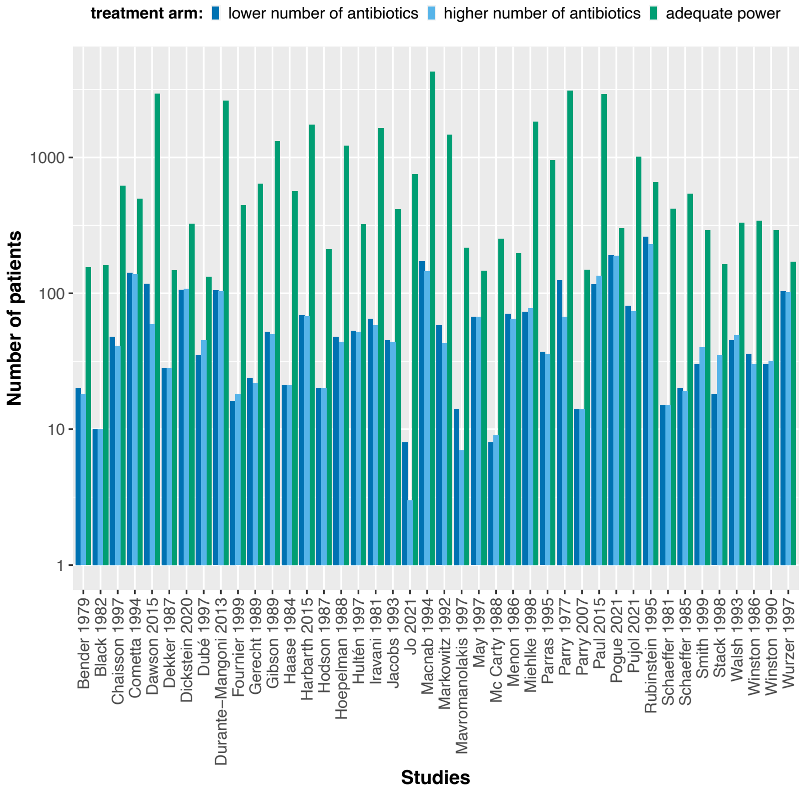


**Fig. S6.** The calculated adequate treatment arm size for each study assuming to detect an odds ratio of 0.5 with 80% power in comparison to the actual treatment arm sizes. The power calculations were performed using the upper confidence interval for the binomial probability of the treatment arm with less antibiotics.

##### *8.2. Trial sequential analysis*

It is expected that pooling data from several RCTs results in a high level of evidence. Nevertheless, meta-analysis might lead to inconclusive results or even misleading ones as meta-analyses can also suffer from low statistical power (69). Therefore, we performed for our two main outcomes a trial sequential analysis (TSA), using the TSA tool version 0.9.5.10 Beta (Copenhagen: The Copenhagen Trial Unit, Centre for Clinical Intervention Research, 2016) to assess how strong and sufficient the evidence of our overall analyses is. For both outcomes the TSA supports that the existing evidence on resistance development is not sufficient and conclusive, as the trial sequential monitoring boundary is not crossed by the Z-curve in any of the cases, nor is the required sample size reached (figure S7). For the TSA calculations we used resistance incidence rate per treatment arm, which we calculated by averaging the incidence rates of all included studies (per outcome). For interested readers technical details of the TSA can be found elsewhere (69, 70). The TSA analysis is an additional analysis, which was not predefined in our study protocol.

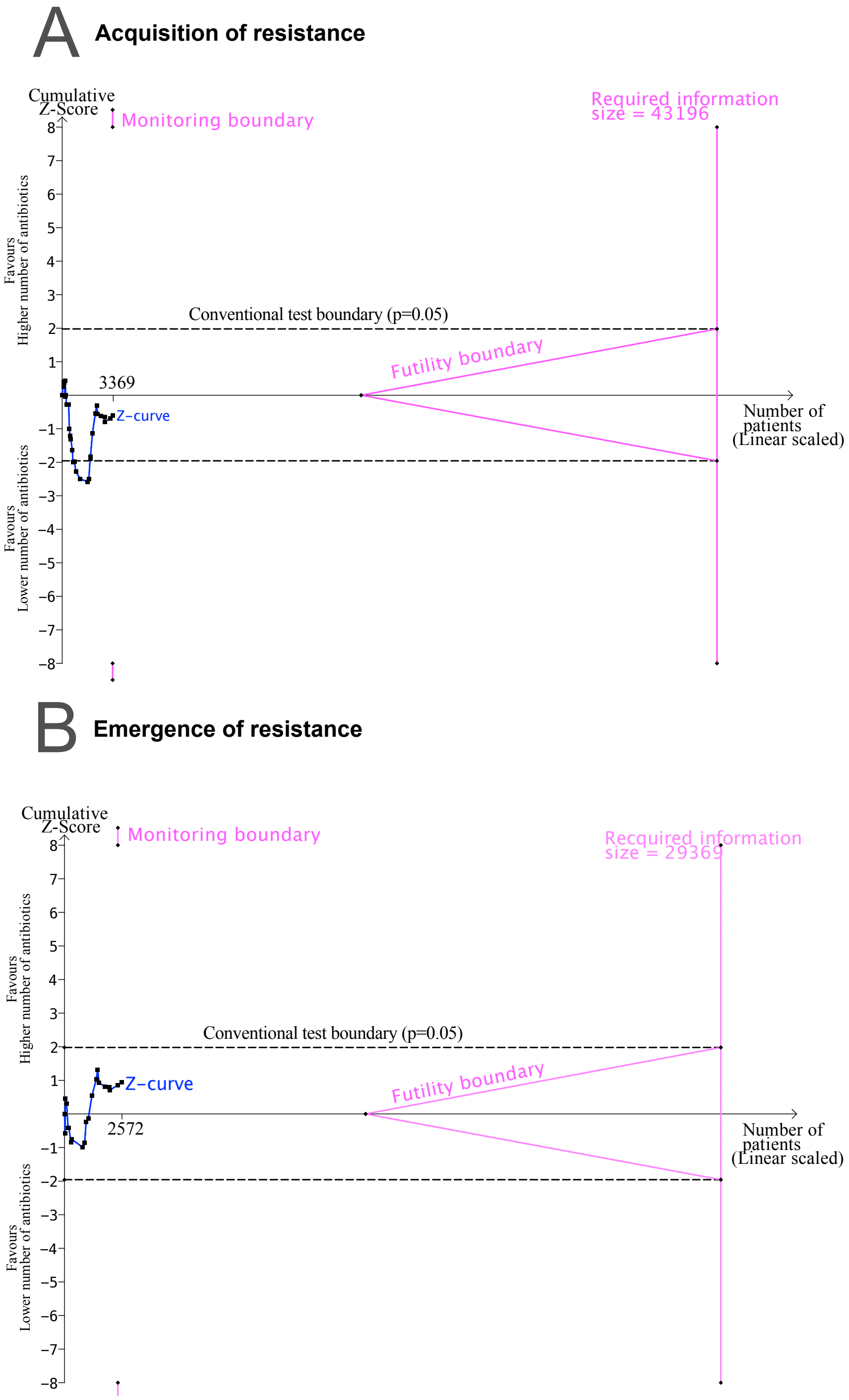


**Fig. S7.** TSA output using 80% power, and 5% significance to detect a relative odds reduction of 50%: A) acquisition of resistance. B) *de novo* emergence of resistance. No sufficient evidence on development of resistance is supported, since the Z-curves do not cross the monitoring nor the futility boundaries, and the required sample size is not reached.

#### 9. Secondary Outcomes

In the evaluation of an optimal antibiotic treatment strategy many factors play a role besides the potential spread of antibiotic resistance and therewith the future potential to treat infections successfully. One important factor, which is naturally the focus of clinical research, is the wellbeing of the patient receiving antibiotic treatment. Antibiotic combination therapy is often associated with a higher medical burden for the treated patient, e.g. through a higher risk of toxicity (71). To present are more comprehensive evaluation of antibiotic combination therapy, we systematically summarised the following outcomes as an indication for the wellbeing of the treated patient: (i) All-cause mortality, (ii) mortality attributable to infection, (iii) treatment failure, (iv) treatment failure due to a change of resistance against the study drugs, and (v) proportion of patients with alterations to the treatment due to adverse events. Additionally, we collected data on acquisition, and *de novo* emergence of resistance against non-administered antibiotics to further assess the risk of resistance spread, which might affect future treatment success. Overall, we did not find any indication of a difference for any of these evaluation metrics of combining a higher number of antibiotics in comparison to less as presented below.

##### *9.1. All-cause mortality*

We extracted the number of patients that died in a study as reported. We did not identify a mortality difference of using a higher number of antibiotics opposed to less (figure S8). One must consider that the estimated pooled OR 0.98 (95% CI 0.79 – 1.21) was based on several RCTs with different sources of potential heterogeneity, which we did not account for in our statistical analysis of secondary outcomes. Nevertheless, the heterogeneity in our random effects model for all-cause mortality could be classified as unimportant ($I^2$=11%). In previously conducted meta-analyses evaluating antibiotic combination therapy mortality was often the main outcome, but the inclusion criteria were less broad, constrained to specific diseases, pathogens, or particular antibiotic combinations. The results of those meta-analyses do not easily generalize to one overall trend, but rather highlight that sub-analyses accounting for specific infections and antibiotic comparisons might be important as we found for our main outcomes of resistance development (72-74). Nevertheless, we found in line with most previous meta-analyses no clear harm or benefit of combining a higher number of antibiotics or less with respect to all-cause mortality.

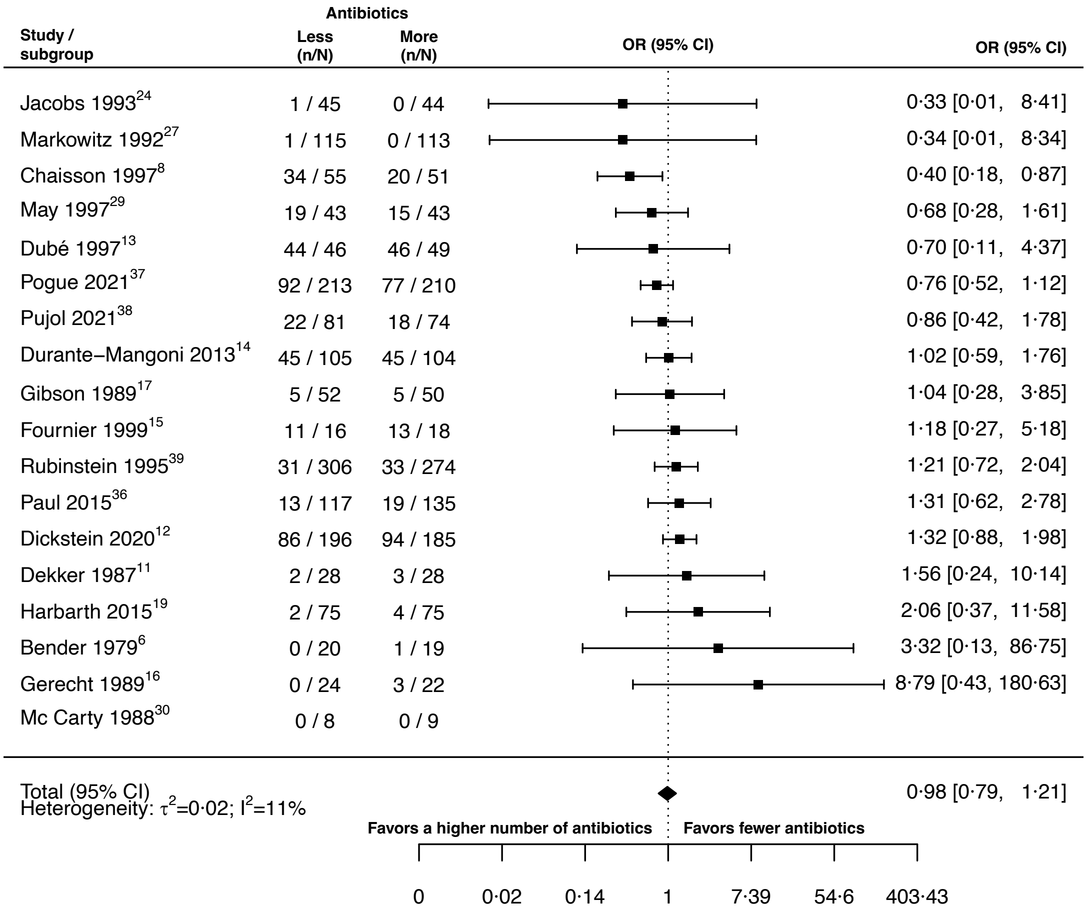


**Fig. S8.** Forest plot of all-cause mortality.

##### *9.2. Mortality attributable to infection*

Besides all cause-mortality we also extracted the number of deaths that the respective study authors attributed to the infection treated. As for all-cause mortality our estimate for mortality attributable to infection indicated no difference between treating with a higher number of antibiotics in comparison to less (pooled OR 1.05, 95% CI 0.64 – 1.71; figure S9), and the model heterogeneity could also be classified as unimportant ($I^2$=12%).

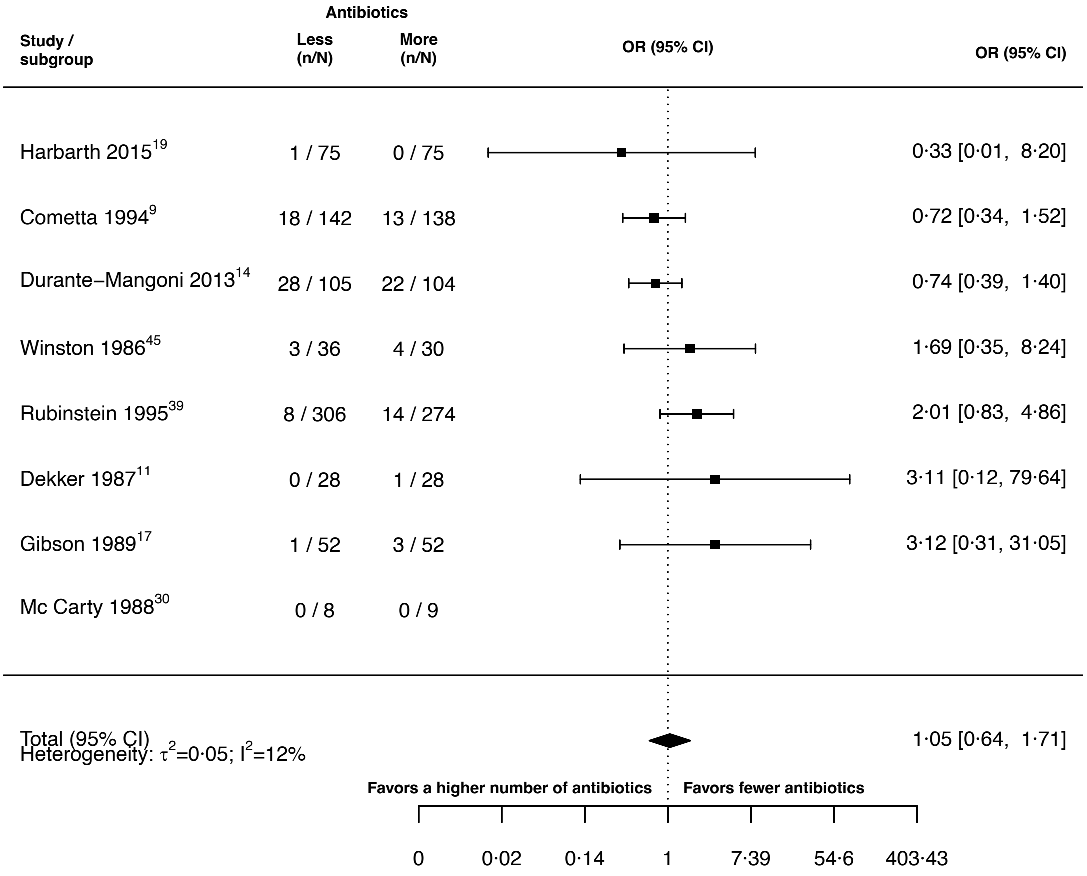


**Fig. S9.** Forest plot of mortality attributable to infection.

##### *9.3. Treatment failure*

We extracted the number of treatment failures in each treatment arm if treatment failure was explicitly defined or classified by the study authors. As the selection of studies for this meta-analysis was not restricted to one specific pathogen, or condition requiring antibiotic treatment, we expected a variety of different reasons for the employment of antibiotics. Out of practicality and to account for the different conditions treated, we decided not to pre-define our own criteria for treatment failure for each condition, but rather use the study's authors interpretation of treatment failure (table S10).

Our estimate gave no indication for a difference in treatment failure when treating with a higher number of antibiotics in comparison with a lower number of antibiotics if treatment failure was considered (pooled OR 0.98, 95% CI 0.66 – 1.47; figure S10). However, we observed a substantial amount of heterogeneity in our model ($I^2$=74%), which might indicate that for some bacterial conditions or some antibiotic combinations there might be a difference.

**Table S10.** Overview of different treatment failure definitions.

| Study | Definition of treatment failure given by the study authors |
|---|---|
| Cometta et al. (1994)**(9)** | Lack of improvement of primary infection, development of a sepsis syndrome or septic shock during treatment, superinfection |
| Durante-Mangoni et al. (2013)**(14)** | No improvement of clinical conditions by day 21 or worsening of the condition at any time, given persistently positive *Acinetobacter baumannii* cultures |
| Gerecht et al. (1989)**(16)** | Continued presence of infecting organism(s) in bile cultures, with persistent indications of cholangitis, or superinfection, or the presence of new infecting organism(s) during or at the end of antibiotic treatment, with indications of cholangitis, or emergence of an infecting organism(s) resistant to gentamicin or mezlocillin during treatment, with indications of cholangitis, or emergence of an infecting organism(s) resistant to gentamicin or mezlocillin during treatment, with indications of cholangitis, or relapse, or recurrence of indications of cholangitis, with the original infecting organism(s) present in cultures of bile or blood within eight weeks after treatment, or death due to uncontrolled infection. |
| Haase et al. (1984)**(18)** | The persisting presence of the pretherapy infecting organism, with or without pyuria, during treatment. |
| Hartbarth et al. (2015)**(19)** | No improvement or worsening in the clinical condition, or a change of the assigned therapy at any time, or death. |
| Jacobs et al. (1993)**(24)** | No apparent response to therapy and no definitive identification of an alternative etiology that would explain this lack of response. |
| Markowitz et al. (1992)**(27)** | Persistence of septic pulmonary emboli, persistence of positive blood or deep tissue cultures, or relapse after the end of presumably adequate treatment. |
| May et al. (1997)**(29)** | Treatment failure was defined as all other situations than success, whereas the primary determinants of success were as follows: patient living, either not fever or a reduction of ≥ 1 °C in initial body temperature, and a blood culture negative for *M. avium* |
| Parry et al. (2007)**(35)** | Continuing fever with at least one other typhoid-related symptom for more than seven days after the start of treatment, or a required change in therapy due to the development of severe complications during treatment (severe gastrointestinal bleeding, intestinal perforation, visible jaundice, myocarditis, pneumonia, renal failure, shock, or an altered conscious level) |
| Paul et al. (2015)**(36)** | Treatment failure at seven days was defined as a composition of death, persistence of fever, persistence of hypotension, non-improving Sequential Organ Failure Assessment score, or persistent bacteraemia on day seven. |
| Pogue et al. (2021)**(37)** | Clinical failure was defined by meeting any of the following criteria: death either during therapy or within 7 days after; receipt of rescue therapy for the trial pathogen within 7 days after treatment, exclusion from the trial due to an adverse event considered related to trial treatment; bacteremia more than 5 days after the begin of therapy for patients with blood stream infections; or failure to improve or worsening of oxygenation by the end of trial treatment in patients with pneumonia. |
| Pujol et al. (2021)**(38)** | No clinical improvement after 3 days of therapy, persistent MRSA bacteraemia at day 7 or later, early discontinuation of therapy due to adverse events or based on clinical judgment, recurrent MRSA bacteraemia before or at test of cure, missing blood cultures at test of cure, and/or death due to any cause before test of cure. |
| Rubinstein et al. (1995)**(39)** | Use of a new antibiotic due to a worsening in clinical condition, isolation of resistant organism, or |

| | |
|---|---|
| | superinfection at the initial site during treatment, no clinical response or death attributed to infection. |

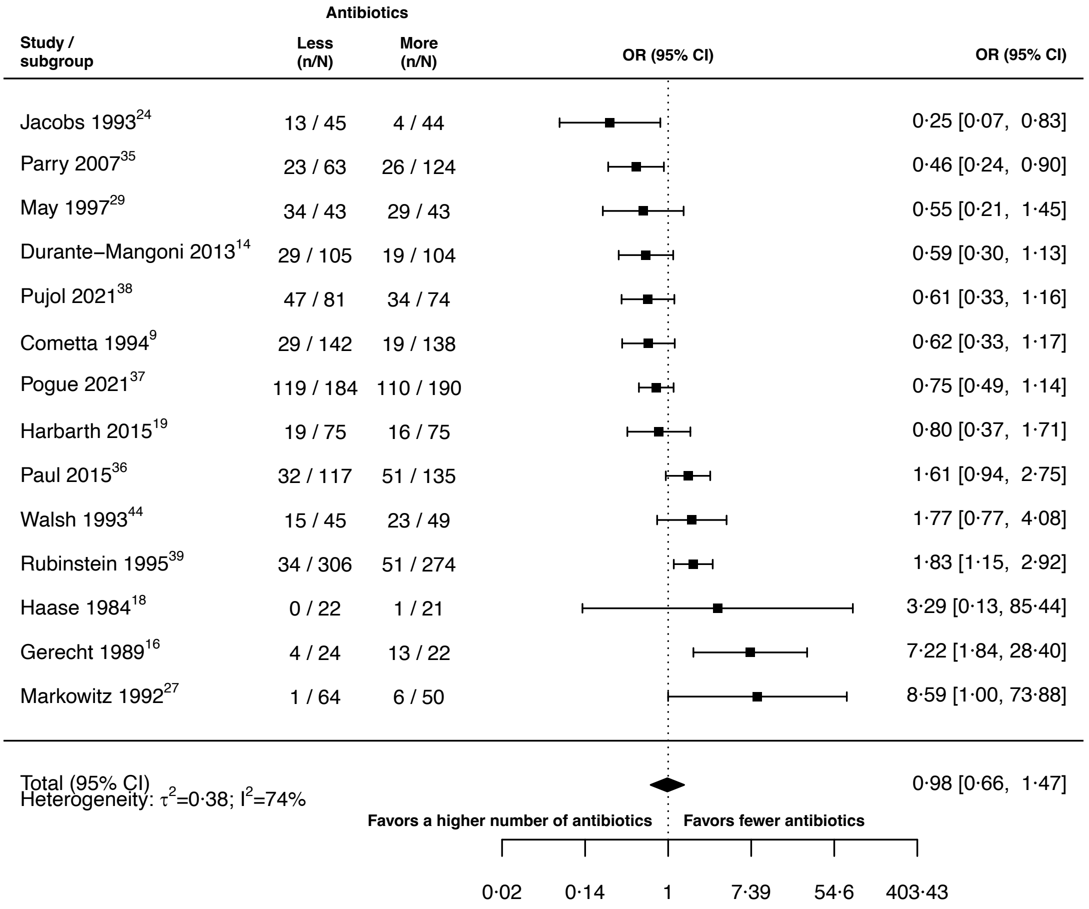


**Fig. S10.** Forest plot of treatment failure.

##### *9.4. Treatment failure due to a change of resistance against the study drugs*

We could only extract information for treatment failure due to a change of resistance against the study drugs from three out of the 42 studies. As one of the studies had zero-events in both treatment arms our statistical summary estimate was only based on two studies and should therefore be interpreted with caution. Nevertheless, as for treatment failure we did not identify a difference of using a higher number of antibiotics in comparison to less when considering treatment failure due to a change of resistance against the study drugs (pooled LOR 0.61, 95% CI 0.29 – 1.28; $I^2$=1%; figure S11).

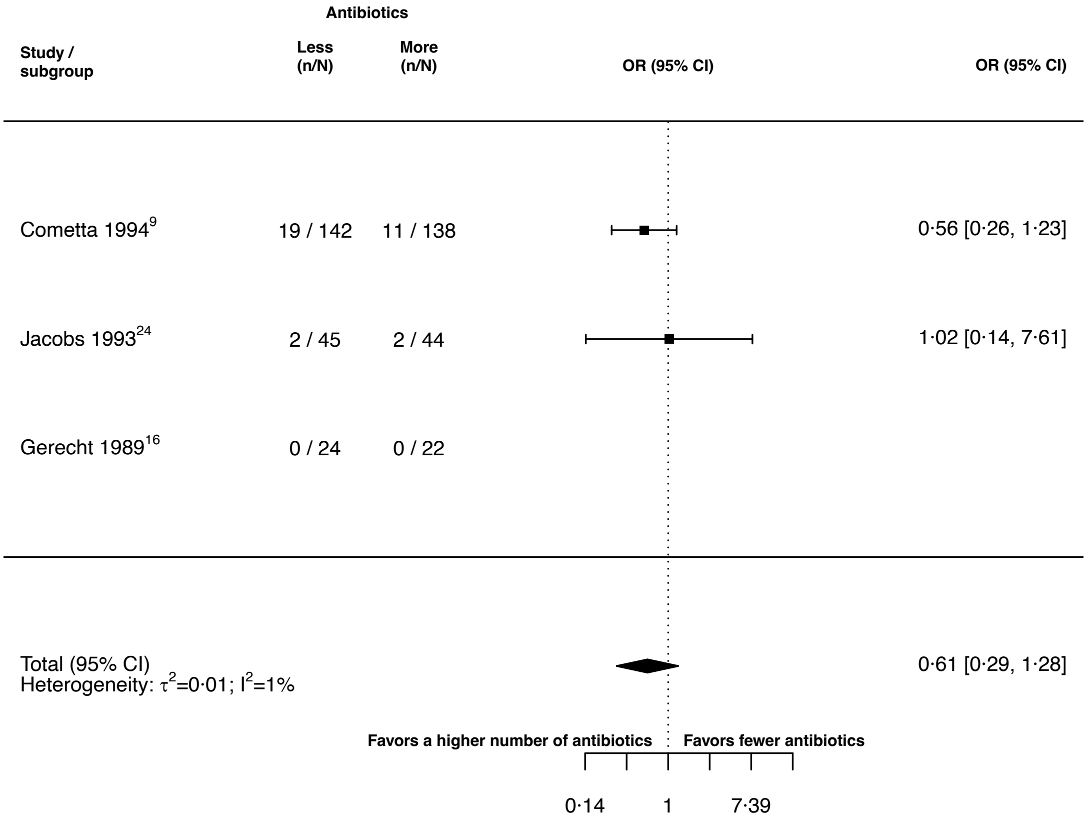


**Fig. S11.** Forest plot of treatment failure due to a change of resistance against the study drugs.

##### *9.5. Alterations of the prescribed treatment due to adverse events*

To get an indication how well the treatments were tolerated by the patients we extracted data on alterations of the prescribed treatment due to adverse events. We did identify benefit of using a lower number of antibiotics in comparison to a higher one. The heterogeneity in the random effects model could be classified as unimportant (pooled OR 1.61, 95% CI 1.12 – 2.31; $I^2$=5%; figure S12).

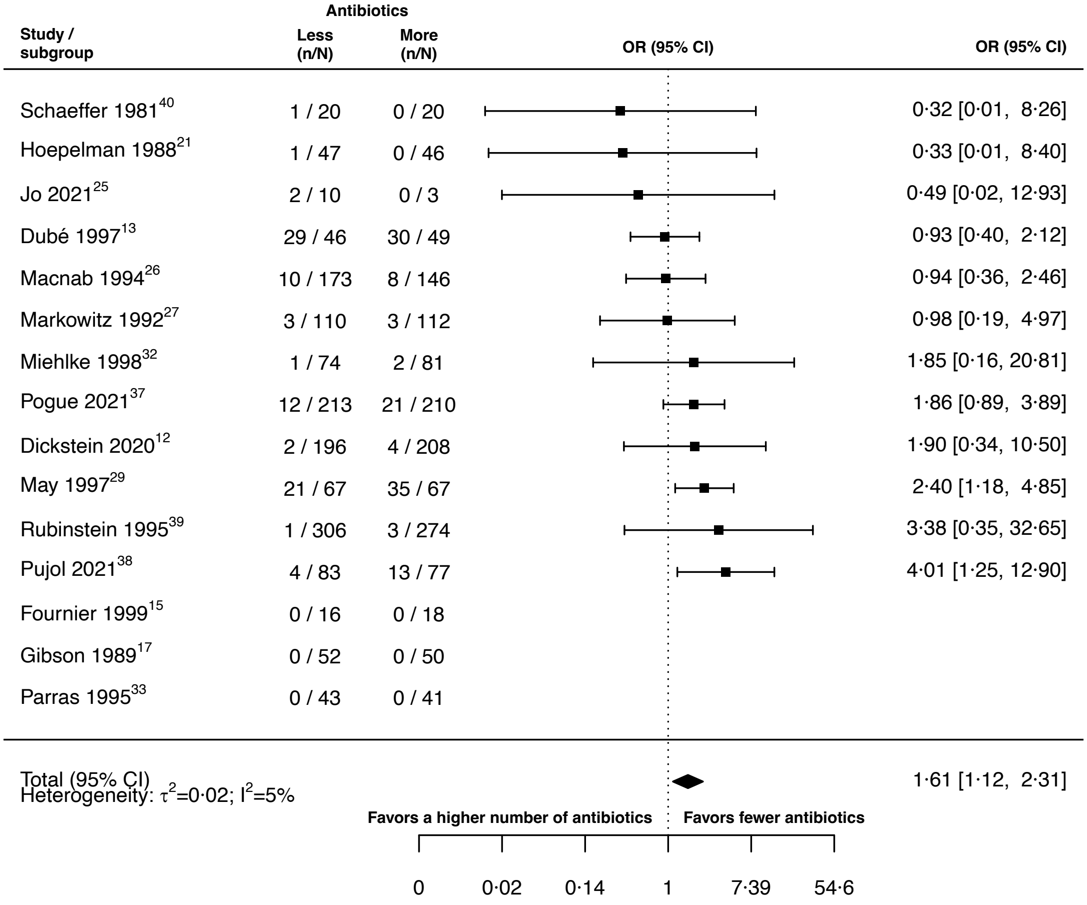


**Fig. S12.** Forest plot of alterations of the prescribed treatment due to adverse events.

##### *9.6. Acquisition of resistance against non-administered antibiotics*

There are several ways of how bacteria may get resistant against antibiotics, one of them is through acquiring antibiotic resistance plasmids. Clinically relevant plasmids often confer resistance against multiple antibiotics (59-61). Therefore, one might expect if a patient is treated with a higher number of antibiotics the chances increase to acquire multidrug resistant plasmids that confer resistances to antibiotics that are not part of the current treatment. In addition, one could expect, that the chances for cross resistances increase, i.e., the obtained resistance confers resistances to several antibiotics, if a higher number of antibiotics is administered. To check this reasoning, we extracted the data for acquisition, and *de novo* emergence of resistance against non-administered antibiotics.

For seven studies we extracted the data for acquisition of resistance against non-administered antibiotics, but we could only use three of them for our statistical analyses as the other studies had zero events in both treatment arms. As the statistical analysis was only based on three studies and the model showed moderate to substantial heterogeneity ($I^2$=60%) our estimate might not be sufficient to confidently give an indication. The pooled LOR of our random effects model suggested no difference in using a higher number of antibiotics in comparison to less to reduce acquisition of resistance against non-administered drugs (OR 0.39, 95% CI 0.02 – 8.48; figure S13).

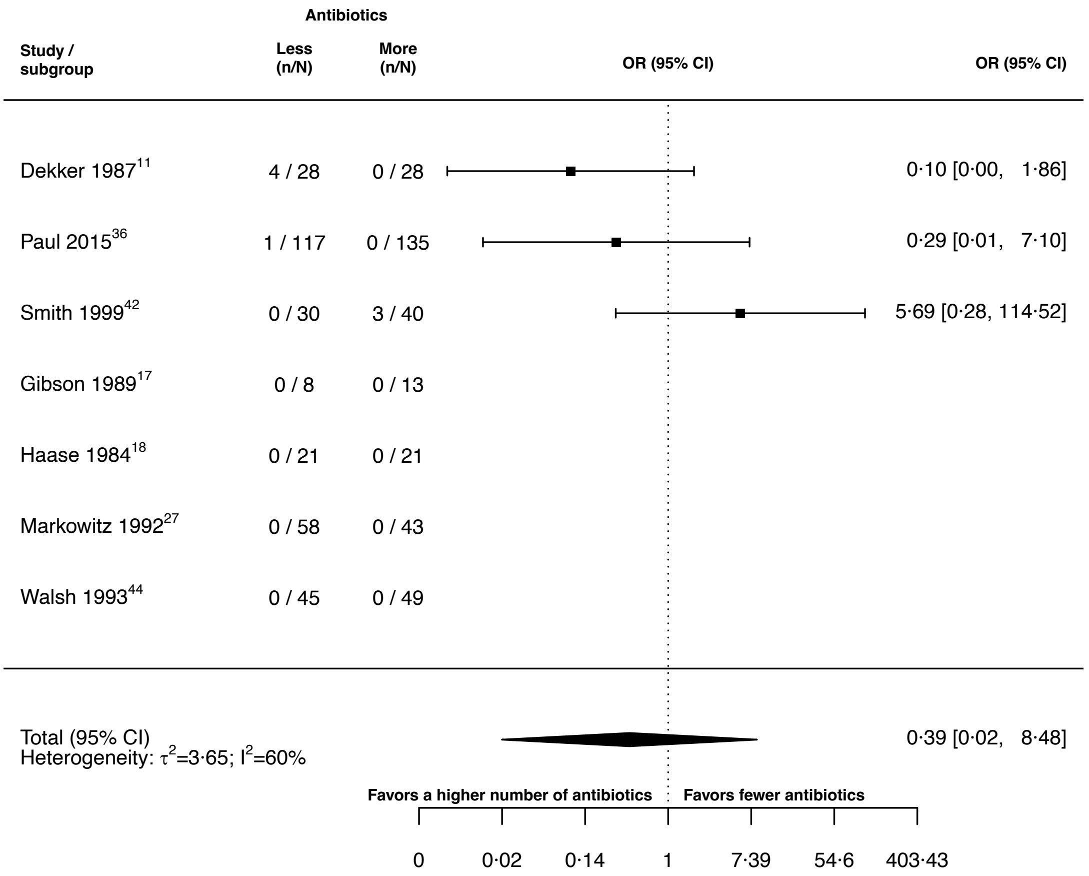


**Fig. S13.** Forest plot of acquisition of resistance against non-administered antibiotics.

##### *9.7.* **De novo** ***emergence of resistance against non-administered antibiotics***

As for the main outcomes we distinguished between acquisition and *de novo* emergence of resistance. According to our definition (main text: Methods), *de novo* emergence of resistance is a subset of acquisition of resistance. For acquisition of resistance against non-administered antibiotics we obtained three studies eligible for the statistical analysis, for *de novo* emergence only two. Therefore, the estimates need to be taken with consideration. As for acquisition of resistance against non-administered antibiotics there was no indication for a difference of using a higher or a lower number of antibiotics (pooled OR 1.91, 95% CI 0.09 – 39.69; $I^2$=33%; figure S14).

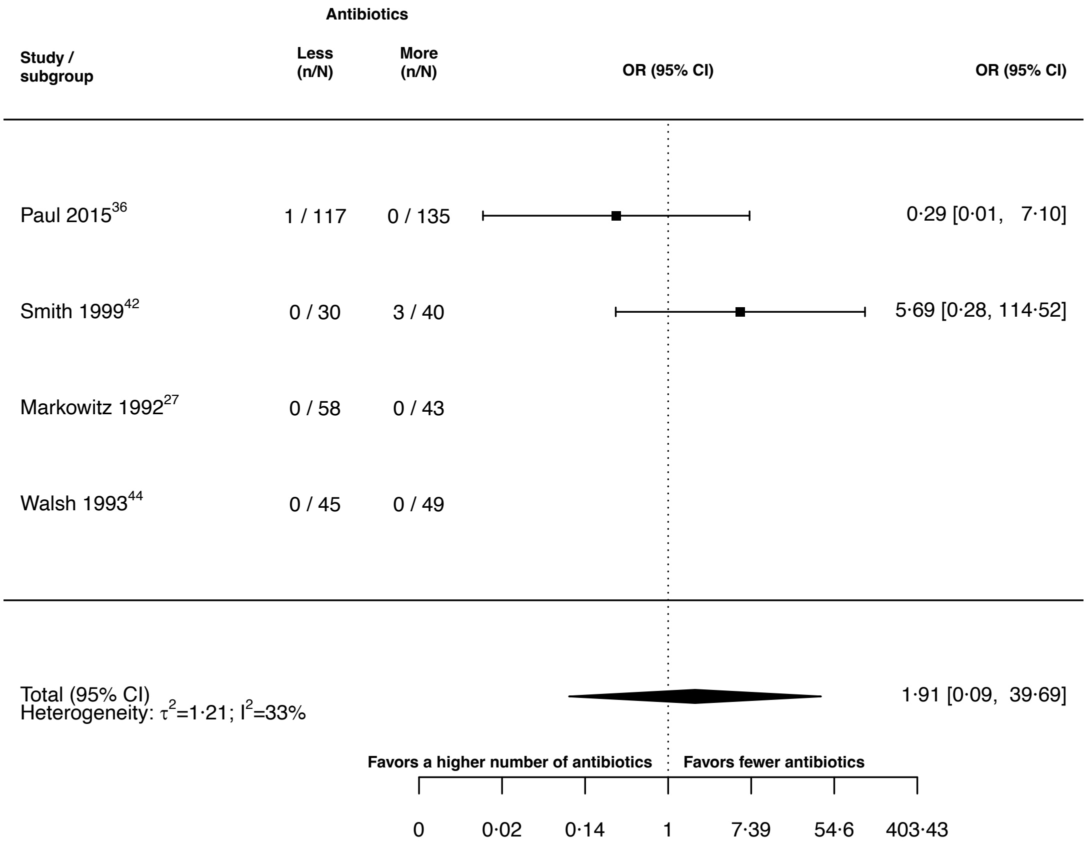


**Fig. S14.** Forest plot of *de novo* emergence of resistance against non-administered antibiotics.

#### 10. List of contacted authors and reasoning for exclusion of studies included in previous meta-analyses

An overview of authors, that were contacted for clarification of study data, is shown in table S11. In our meta-analysis we excluded some studies that were included in previous meta-analyses focusing on resistance development (72, 75). An overview of those studies and an exclusion reason is given in table S12.

**Table S11.** List of studies for which study authors or institutions were contacted. An indication is given whether clarifying information was obtained.

| Study | Person/Institution contacted | Information sufficient for paper inclusion obtained (yes/no) |
|---|---|---|
| Bazolli 1998**(76)** | Franco Bazolli | no |
| Benson 2000**(77)** | Constance Benson | no |

| | | |
|---|---|---|
| Bochenek 2003**(78)** | David Yates Graham; Wieslaw Bochenek | no |
| Bosso 1988**(79)** | John Bosso | no |
| Bow 1987**(80)** | Eric Bow | no |
| Cruciani 1989**(81)** | Mario Cruciani | no |
| Dalgic 2013 **(82)** | Nazan Dalgic | no |
| De Pauw 1985**(83)** | Ben de Pauw | no |
| De Pauw 1987**(84)** | Ben de Pauw | no |
| Dinubile 2005**(85)** | Mark Dinubile | no |
| East African/British medical research councils 1972**(86)** | Research office of the royal Brompton & Harefield hospitals | no |
| Frank 2002**(87)** | Elliot Frank | no |
| Gold 1985**(88)** | Ronald Gold | no |
| Grossman 1994**(89)** | Ronald Grossman | no |
| Grabe 1986**(90)** | Magnus Grabe | no |
| Guerrant 1981**(91)** | Richard Guerrant | no |
| Heyland 2008**(92)** | Daren Heyland | no |
| Hodson 1987**(20)** | Margaret Hodson | no |
| Hoepelman 1988**(21)** | Andy I.M. Hoepelman | no |
| Jackson 1986**(93)** | Mary Anne Jackson | no |
| Liang 1990**(94)** | Raymond Hin Suen Liang | no |
| McLaughlin 1983**(95)** | John McLaughlin | no |
| Muder 1994**(96)** | Robert Muder | no |
| Padoan 1987**(97)** | Rita Padoan | no |
| Paul 2015**(36)** | Mical Paul | yes |
| Parry 2007**(35)** | Christopher Parry | yes |
| Pujol 2021**(38)** | Miquel Pujol | yes |
| Schaad 1997**(98)** | Urs Schaad | no |
| Shawky 2022**(99)** | Sherief Bad-Elsalam | no |
| Sun 2022**(100)** | Jia Fan | no |

**Table S12.** Table of studies, which were included in previous meta-analyses, but excluded in our study. The reason for exclusion is indicated. *In our protocol we stated, that we would include articles in Russian language. However, since VNK, the only Russian speaking author, did not screen all the papers from our systematic search for inclusion, we excluded studies in Russian language.

| **Study** | **Inclusion in previous meta-analyses** | **Reason for exclusion** | **Identified with our search strategy** |
|---|---|---|---|
| Carbon 1987**(101)** | Paul 2014(72) | Not accessible via ETH Zurich library services | no |
| Cone 1985**(102)** | Bliziotis 2005(75) | No data on resistance emergence, due to no clear statement how many resistances are measured in the treatment arm with more antibiotics | no |
| Croce 1993**(103)** | Bliziotis 2005(75) | No proper randomisation of treatment strategies, i.e., the trial was conducted in different phases | yes |
| Gribble 1983**(104)** | Bliziotis 2005(75) | No fixed treatment, since antibiotics could be substituted during treatment | yes |
| Iakovlev 1998**(105)** | Paul 2014(72) | Russian language* | no |
| Klatersky 1973**(106)** | Paul 2014(72) | Not clearly extractable how many patients developed resistance | no |
| Mandell 1987**(107)** | Bliziotis 2005(75), Paul 2014(72) | Treatment is not fixed due to alterations of treatment based on the infecting organism | no |
| Sculier 1982**(108)** | Paul 2014(72) | No proper comparison, since the study does not compare per se a different number of antibiotics but | no |

| | | | |
|---|---|---|---|
| | | adds an additional way of administration of the same antibiotic | |
| German and Austrian Imipenem/Cilastatin study group 1992**(109)** | Bliziotis 2005(75), Paul 2014(72) | No fixed treatment, as an additional antibiotic was allowed to be administered only in the treatment arm with more antibiotics | no |

#### 11. Search strategy

##### *11.1. PubMed*

((((((((((((("Bacterial Infections/Drug Therapy"[mesh]) OR "Bacterial Infections/drug effects"[Mesh]) OR "Bacteria/drug effects"[Mesh]) OR "Bacteria/Drug Therapy"[mesh]) OR (((infection[tiab] OR infections[tiab]) AND bacteria*)))) AND (((((((((((((((((("beta-Lactams/Administration and Dosage"[mesh] OR "beta- Lactams/Therapeutic Use"[mesh])) OR ("Aminoglycosides/Administration and Dosage"[mesh] OR "Aminoglycosides/Therapeutic Use"[mesh])) OR ("Chloramphenicol/Administration and Dosage"[mesh] OR "Chloramphenicol/Therapeutic Use"[mesh])) OR ("Glycopeptides/Administration and Dosage"[mesh] OR "Glycopeptides/Therapeutic Use"[mesh])) OR ("Rifamycins/Administration and Dosage"[mesh] OR "Rifamycins/Therapeutic Use"[mesh])) OR ("Streptogramins/Administration and Dosage"[mesh] OR "Streptogramins/Therapeutic Use"[mesh])) OR ("Sulfonamides/Administration and Dosage"[mesh] OR "Sulfonamides/Therapeutic Use"[mesh])) OR ("Tetracyclines/Administration and Dosage"[mesh] OR "Tetracyclines/Therapeutic Use"[mesh])) OR ("Macrolides/Administration and Dosage"[mesh] OR "Macrolides/Therapeutic Use"[mesh])) OR ("Oxazolidinones/Administration and Dosage"[mesh] OR "Oxazolidinones/Therapeutic Use"[mesh])) OR ("QUINOLONES/Administration and Dosage"[mesh] OR "QUINOLONES/Therapeutic Use"[mesh])) OR ("Lipopeptides/Administration and Dosage"[mesh] OR "Lipopeptides/Therapeutic Use"[mesh])) OR ("Anti-Bacterial Agents/Administration and Dosage"[mesh:noexp]))) OR "Anti-Bacterial Agents/Therapeutic Use"[mesh:noexp]) OR "Anti-Bacterial Agents/Therapy"[mesh:noexp]) OR antibiotic*[tiab])) AND (((((((("Drug Therapy, Combination"[mesh:noexp]) OR "drug combinations"[mesh:noexp]) OR "trimethoprim, sulfamethoxazole drug combination"[mesh:noexp]) OR "Drug Synergism"[mesh:noexp]))) OR (combination[tiab] AND (therapy[tiab] OR therapies[tiab]))) OR combinationtherap*[tiab])) AND ((("Drug Resistance, Bacterial"[Mesh]) OR "Drug Resistance, Microbial"[Mesh:noexp]) OR resistan*[tiab]))) NOT (((("Complementary Therapies"[Mesh]) OR "Plant Extracts"[Mesh]) OR bismuth[tiab]) OR "Bismuth"[Mesh]))) AND "Controlled Clinical Trial"[Publication Type]

##### *11.2. CENTRAL*

#1 MeSH descriptor: [Bacterial Infections] explode all trees and with qualifier(s): [drug therapy - DT]
#2 MeSH descriptor: [Bacteria] explode all trees and with qualifier(s): [drug effects -
#3 ((infection):ti,ab,kw OR (infections):ti,ab,kw) AND bacteria*
#4 MeSH descriptor: [beta-Lactams] explode all trees and with qualifier(s): [administration & dosage - AD,
therapeutic use - TU]
#5 MeSH descriptor: [Chloramphenicol] explode all trees and with qualifier(s): [administration & dosage - AD, therapeutic use - TU]
#6 MeSH descriptor: [Aminoglycosides] explode all trees and with qualifier(s): [administration & dosage - AD, therapeutic use - TU]
#7 MeSH descriptor: [Glycopeptides] explode all trees and with qualifier(s): [administration & dosage - AD, therapeutic use - TU]
#8 MeSH descriptor: [Rifamycins] explode all trees and with qualifier(s): [administration & dosage - AD, therapeutic use - TU]
#9 MeSH descriptor: [Streptogramins] explode all trees and with qualifier(s): [administration & dosage - AD, therapeutic use - TU]

#10 MeSH descriptor: [Sulfonamides] explode all trees and with qualifier(s): [administration & dosage - AD, therapeutic use - TU]
#11 MeSH descriptor: [Macrolides] explode all trees and with qualifier(s): [administration & dosage - AD, therapeutic use - TU]
#12 MeSH descriptor: [Tetracyclines] explode all trees and with qualifier(s): [administration & dosage - AD, therapeutic use - TU]
#13 MeSH descriptor: [Oxazolidinones] explode all trees and with qualifier(s): [administration & dosage - AD, therapeutic use - TU]
#14 MeSH descriptor: [Quinolones] explode all trees and with qualifier(s): [administration & dosage - AD, therapeutic use - TU]
#15 MeSH descriptor: [Lipopeptides] explode all trees and with qualifier(s): [administration & dosage - AD, therapeutic use - TU]
#16 MeSH descriptor: [Anti-Bacterial Agents] this term only and with qualifier(s): [administration & dosage - AD, therapeutic use - TU]
#17 (antibiotic*):ti,ab,kw
#18 MeSH descriptor: [Drug Therapy, Combination] this term only
#19 MeSH descriptor: [Drug Combinations] this term only
#20 MeSH descriptor: [Trimethoprim, Sulfamethoxazole Drug Combination] this term only
#21 MeSH descriptor: [Drug Synergism] this term only
#22 ((combination):ti,kw,ab) NEAR/3 ((therapy):ti,kw,ab OR (therapies):ti,ab,kw)
#23 (combinationtherap*):ti,ab,kw
#24 MeSH descriptor: [Drug Resistance, Bacterial] explode all trees
#25 MeSH descriptor: [Drug Resistance, Microbial] this term only
#26 (resistan*):ti,ab,kw
#27 MeSH descriptor: [Complementary Therapies] explode all trees
#28 MeSH descriptor: [Plant Extracts] explode all trees
#29 (bismuth):ti,ab,kw
#30 MeSH descriptor: [Bismuth] explode all trees
#31 {OR # 1-# 3}
#32 {OR # 4-#17}
#33 {OR #18-#23}
#34 {OR #24-#26}
#35 {AND #31-#34}
#36 {OR # 27-#30}
#37 #35 NOT #36

##### *11.3. EMBASE*

#26. #24 AND #25
#25. 'controlled clinical trial'/exp
#24. #23 NOT #22
#23. #18 AND #19 AND #20 AND #21
#22. #14 OR #15 OR #16 OR #17
#21. #12 OR #13
#20. #7 OR #8 OR #9 OR #10 OR #11
#19. #5 OR #6
#18. #1 OR #2 OR #3 OR #4
#17. 'herbal medicine'/exp
#16. 'alternative medicine'/exp
#15. 'bismuth'/exp
#14. bismuth:ti,ab,kw
#13. resistan*:ti,ab,kw
#12. 'antibiotic sensitivity'/exp
#11. (combination NEAR/3 (therapy OR therapies)):ti,ab,kw
#10. combinationtherap*:ti,ab,kw
#9. 'antibiotic agent'/exp/dd_cb
#8. 'drug potentiation'/de
#7. 'combination drug therapy'/de

#6. 'antibiotic*':ti,ab,kw
#5. 'antibiotic agent'/exp
#4. (infection:ti,ab,kw OR infections:ti,ab,kw) AND bacteria*
#3. 'bacterial infection'/exp
#2. 'bacterium'/exp
#1. 'prokaryotes by outer appearance'/exp

##### *11.4. Screening of eligible trials and previous meta-analyses*

In addition to the systematic database search, we also screened the references of eligible studies and the trials included in two previous meta-analyses (72, 75). With the database search we identified 41 studies. While screening the references of those 41 studies we identified one additional study (45), which meets our inclusion criteria. This additional study was not identified in our search strategy as neither the abstract nor database specific identifiers gave any indication that resistance was measured in this study. The screening of the trials included in two previous analyses did not result in inclusion of further studies (table S12).
